# A functional genomic framework to elucidate novel causal non-alcoholic fatty liver disease genes

**DOI:** 10.1101/2024.02.03.24302258

**Authors:** Peter Saliba-Gustafsson, Johanne M. Justesen, Amanda Ranta, Disha Sharma, Ewa Bielczyk-Maczynska, Jiehan Li, Laeya A. Najmi, Maider Apodaka, Patricia Aspichueta, Hanna M. Björck, Per Eriksson, Anders Franco-Cereceda, Mike Gloudemans, Endrina Mujica, Marcel den Hoed, Themistocles L. Assimes, Thomas Quertermous, Ivan Carcamo-Orive, Chong Y. Park, Joshua W. Knowles

**Affiliations:** Department of Medicine, Division of Cardiovascular Medicine and Cardiovascular Institute, Stanford University, Stanford, CA, USA; CardioMetabolic Unit at the Department of Medicine, Huddinge, Karolinska Institutet, Stockholm, Sweden; Stanford Diabetes Research Center, Stanford, CA, USA; Stanford Cardiovascular Institute, Stanford University School of Medicine, CA, USA; Novo Nordisk Foundation Center for Basic Metabolic Research, University of Copenhagen, Denmark; The Hormel Institute, University of Minnesota, MN, USA; University of the Basque Country (UPV/EHU), Faculty of Medicine and Nursing, Department of Physiology, Leioa, Spain; National Institute for the Study of Liver and Gastrointestinal Diseases (CIBERehd, Instituto de Salud Carlos III); Division of Cardiovascular Medicine, Centre for Molecular Medicine, Department of Medicine, Solna, Karolinska Inistitutet, Stockholm, Karolinska University Hospital, Solna, Sweden; Department of Molecular Medicine and Surgery, Karolinska Inistitutet, Stockholm, Sweden; Department of Pathology, Stanford University School of Medicine, CA, USA; Department of Immunology, Genetics and Pathology, Uppsala University, Sweden; VA Palo Alto Health Care System, Palo Alto CA, USA; IKERBASQUE, Basque Foundation for Science, Bilbao, Spain; Stanford Prevention Research Center, Stanford, CA, USA

**Keywords:** NAFLD, CRISPR interference, genetic epidemiology, Perturb-seq, HepaRG cells

## Abstract

**Background & Aims:** Non-alcoholic fatty liver disease (NAFLD) is the most prevalent chronic liver pathology in western countries, with serious public health consequences. Efforts to identify causal genes for NAFLD have been hampered by the relative paucity of human data from gold-standard magnetic resonance quantification of hepatic fat. To overcome insufficient sample size, genome-wide association studies using NAFLD surrogate phenotypes have been used, but only a small number of loci have been identified to date. In this study, we combined GWAS of NAFLD composite surrogate phenotypes with genetic colocalization studies followed by functional in vitro screens to identify bona fide causal genes for NAFLD.

**Approach & Results:** We used the UK Biobank to explore the associations of our novel NAFLD score, and genetic colocalization to prioritize putative causal genes for *in vitro* validation. We created a functional genomic framework to study NAFLD genes *in vitro* using CRISPRi. Our data identify *VKORC1, TNKS, LYPLAL1* and *GPAM* as regulators of lipid accumulation in hepatocytes and suggest the involvement of *VKORC1* in the lipid storage related to the development of NAFLD.

**Conclusions:** Complementary genetic and genomic approaches are useful for the identification of NAFLD genes. Our data supports *VKORC1* as a *bona fide* NAFLD gene. We have established a functional genomic framework to study at scale putative novel NAFLD genes from human genetic association studies.

## Introduction

Non-alcoholic fatty liver disease (NAFLD) is the most common chronic liver condition, with serious public health consequences. Globally, at least 25% of adults are estimated to suffer from NAFLD, and cardiovascular disease is the leading cause of death among these patients. (1, 2) NAFLD displays a wide spectrum of liver pathology, ranging from nonalcoholic fatty liver, which is typically benign, to non-alcoholic steatohepatitis (NASH), characterized by steatosis and features of cellular injury, such as inflammation and hepatocyte ballooning. NASH may progress to liver cirrhosis, hepatic failure, and hepatocellular carcinoma in the absence of significant alcohol consumption. The degree of steatosis can be measured through various imaging techniques but the gold standard of these is abdominal magnetic resonance imaging (MRI). However, abdominal MRI is not typically conducted on asymptomatic individuals, often leaving NAFLD undiagnosed for years.

Genome-wide association studies (GWAS) have been used to identify associations between NAFLD and common genetic variants. Due to the scarcity of MRI data, identifying risk loci for NAFLD has been slower than for other cardiometabolic diseases or their risk factors (e.g., body mass index (BMI)) or biochemical measures (e.g., serum liver enzymes and lipids levels), and other complex cardiometabolic diseases such as obesity, and diabetes. One way to overcome the challenge of data scarcity in NAFLD is to comprise latent proxies for NAFLD using data that is more readily available in large cohort studies. For instance, Bedogni et al. (3) established the fatty liver index (FLI) as a surrogate variable for NAFLD; however, FLI did not outperform waist circumference in predicting NAFLD in a validation study. (4)

Aiming to increase our understanding of the molecular etiology of NAFLD, we here generate (NAFLD-S), a composite variable of anthropometric and biochemical variables to predict liver fat. By using an alternative surrogate to predict liver fat, and running GWAS combined with genetic colocalization, we identify novel loci associated with NAFLD. The use of genetic colocalization aids in inferring causality and serves to prioritize genes for functional follow-up. We use CRISPR-interference (CRISPRi) to interrogate the impact of multiple genes on both transcriptional changes and functional phenotypes, at a single-cell level. (5–9) We characterize a subset of putative NAFLD genes *in vitro* and *in vivo* through an integrated framework and identify *VKORC1* as a likely causal NAFLD gene.

## Experimental procedures

### Study population

This research was conducted using UK Biobank (UKB) data under application number 13721. The UKB is a cohort of over 500,000 adults that has tracked health behaviors, anthropometric measurements, and medical history. Biological samples that have been acquired longitudinally since the subjects’ enrollment in 2006-2010. A subset of white British participants in the UKB was utilized in the current study, a sub-population which has been described in detail (10). Briefly, we excluded individuals who withdrew consent (n=167), and those who reported excessive alcohol consumption (n=128,477), which was defined as weekly alcohol consumption of ≥140 grams for women and ≥210 grams for men as per prevailing European guidelines (11). Further, we excluded those with other known liver diseases, alcohol use disorder, and HIV infection based on ICD-9 and ICD-10 codes (n=3,022). Individuals with short-term poor prognosis including diagnosis of metastatic cancer within one year of the baseline visit and palliative care or hospice status based on ICD-9 and ICD-10 codes were also excluded (n=4,497). After exclusions 242,524 individuals remained, whose characteristics have been described (10). The UKB study was approved by the Northwest Multi-Center Research Ethics Committee and all participants provided written informed consent to participate. The UKB study protocol is available online (12).

### Generating the NAFLD score in UKB

In the UKB, true NAFLD cases were first identified using existing data on liver fat from MRI (data fields 22402, 22436 and 24352). Individuals with a liver fat percent ≥ 5.5% were considered to have NAFLD, which resulted in a population of 2,544 NAFLD cases and 10,168 controls. The 5.5% cut-off is higher than the 5% convention, but motivated to reduce the risk of measurement error impacting the results. We utilized biochemical and anthropometric data to determine a NAFLD score in the full subset of 242,524 participants in the UKB. Briefly, we performed a multivariate logistic regression analysis to estimate effect sizes of the biochemical and anthropometric predictors of liver fat percentage from MRI data. ***Table 1*** shows the variables included in the prediction models and their effect sizes. We constructed a NAFLD score for the subset of UKB, as well as calucated the fatty liver index (FLI) (3), the difference lies in that the NAFLD score includes more predictors of liver fat from MRI than the FLI (see equation 1 below). The score was derived by multiplying the value for each biochemical and anthropometric trait with its effect size (beta), and then rounded to the nearest integer between 0 and 100 (Equation 1). The NAFLD score was compared with FLI, alanine aminotransferase (ALT), triglycerides (TG), gamma-glutamyl transpeptidase (GGT) and BMI in predicting liver fat percentage using a receiver operating characteristic (ROC) curve. Ultimately, the NAFLD score was used as the surrogate for NAFLD to conduct a GWAS in 242,524 participants in the UKB.

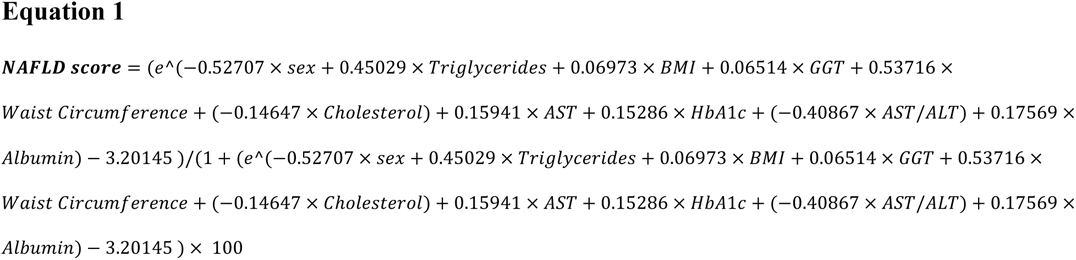

### Genome-wide association analyses

Genome-wide association analyses (GWAS) were carried out using both the generated NAFLD score as a continuous variable (NAFLD-S), ALT, and NAFLD status from MRI (MRI_UKB) in 242,524 white Brits with an at most moderate alcohol consumption. Analyses were carried out in Plink 2, and were adjusted for age, sex, genotyping batch and the first 10 genetic principal components. Imputed SNPs with INFO<0.8 were excluded, and dependent SNPs were pruned out using a 50kb window and a *R^2^* threshold of 0.8.

### Genetic colocalization analyses

SNPs associated with either cALT (chronic ALT), ALT (one point measurement), ALP, GGT, NAFLD score or NAFLD status from MRI were tested for genetic colocalization using the GTEx (v8) eQTL and sQTL data in metabolically relevant tissues (liver, subcutaneous adipose tissue, visceral adipose tissue, skeletal muscle and pancreas). Briefly, summary statistics from previously conducted GWASes (cALT, ALP, GGT) were obtained, and integrated with the GWASes conducted herein (NAFLD-S, ALT and NAFLD status). The colocalization analyses compute the probability that genetic association signals for trait and a QTL feature are produced by a common causal variant, as well as gene-level colocalizations. We used a novel, in-house, custom integration of the FINEMAP (13) and eCAVIAR (14) methods, which has been described in detail (15), to calculate gene-level colocalization scores. Genome-wide significance threshold was determined as 5e-8 in all except the two MRI GWASes where the significance threshold was set at 1e-5. QTL threshold was set at 1e-5. A colocalization score of 0.35 or above was considered as a coloc. All colocalizations were then investigated for gene set enrichment using GSEA (http://www.gsea-msigdb.org/gsea/index.jsp), to elucidate pathways that are disturbed in NAFLD.

### Cellular model

The HepaRG™ cell line (Sigma Aldrich) was established from a tumor of a female patient suffering from chronic hepatitis C (HVC) infection and hepatocarcinoma. HepaRG™ cells do not contain any part of the HCV genome nor express any HCV protein. (16–18). *In vitro*, maximum cell differentiation is reached when cells are exposed to 2 weeks of differentiation media 14 days after seeding, and 40 to 50% of the confluent cell population is hepatocyte-like in nature, with a morphology close to that of primary human hepatocytes (PHHs). A genome-wide gene expression profile analysis showed that for most genes encoding phase 1 and 2 drug metabolizing enzymes and drug transporters, the differences between HepaRG™ cells and PHHs were much smaller than between HepG2 cells and PHHs. (19)

### Generation, characterization, and validation of a HepaRG cell line suitable for gene editing

HepaRG cells were transduced with lentiviral vectors carrying the pHR-SFFV-KRAB-dCas9-P2A-mCherry plasmid (Addgene, #60954), which was a gift from Jonathan Weissman. (20). Lentivirus was produced as previously described. (21) Transduced cells were selected based on mCherry expression using FACS. The resulting KRAB-dCas9-mCherry cell line was characterized by single-cell sequencing before and after differentiation using the Chromium Next GEM Single Cell 3’ GEM, Library & Gel Bead Kit (10X Genomics). Knockdown efficiency of KRAB-dCas9 was tested by transduction of single guide RNAs (individual sgRNAs inserted into pBA904 (Addgene #122238)) targeting *PNPLA3*, followed by cell sorting and RT-qPCR.

### sgRNA cloning and Perturb-seq library preparation

Two to three sgRNAs targeting each gene of interest, and five control non-targeting sgRNAs, were chosen from the Weissman human genome-wide CRISPRi-v2 library (22). The top and bottom insert oligos were ordered as single stranded DNA (ssDNA) from Integrated DNA Technologies. They consisted of the sgRNA sequence (reverse-compliment for bottom) with the following overhangs: TGG (5’) and GTTTAAGAGC (3’) (top), and TTAGCTCTTAAAC (5’) and CAACAAG (3’) (bottom). The 3’ direct-capture Perturb-seq plasmid (pBA904, Addgene #122238) was digested with BstXI and BlpI, and purified from agarose gel using QIAquick Gel Extraction Kit (QIAGEN). Top and bottom sgRNA DNA oligos were annealed, ligated into the digested 3’ direct-capture Perturb-seq backbone, and transformed into NEB Stable *E.coli*. Plasmid DNA was prepared from liquid cultures originating from single colonies and sgRNA sequences were confirmed using Sanger sequencing. The plasmids were combined at equal molar ratios to a plasmid library.

### Lentivirus packing and transduction

Lentiviral stock of the sgRNA plasmid library was produced as previously described (21). Breifly, the sgRNA plasmid library was co-transfected with lentiviral packaging plasmids pMD2.G and pCMVdR8.91 in HEK293T cells, and levtiviral supernatant was collected 48 hours later. To establish a multiplicity of infection (MOI) of 0.10-0.15, HepaRG cells were transduced with serial dilutions of lentiviral stock in the presence of 8 ug/ml polybrene, cultured for two days, and analyzed for blue fluorescent protein (BFP) using flow cytometry.

### CRISPRi screens and Perturb-seq experiments

On the day of the single-cell capture (day 42 of HepaRG cell culturing), cells were trypsinized, strained, and stained with the live/dead stain SYTOX Green Ready Flow Reagent (Thermo Fisher), according to the manufacturer’s protocol. Cells were kept on ice throughout. Live (SYTOX-negative), BFP/mCherry^+/+^ single cells were isolated using FACS at the Stanford Shared FACS Facility. Next, live, gene-edited cells underwent microfluidic single-cell capture on the 10X Chromium Controller device at the Stanford Genomics Service Center. In brief, single cells were encapsulated with individual Gel Beads-in-emulsion (GEMs) using the Chromium Next GEM Single Cell 3’ GEM, Library & Gel Bead Kit with the CRISPR feature barcodes technology (10X Genomics). In-drop reverse transcription and cDNA amplification were conducted according to the manufacturer’s protocol to construct expression and feature barcode libraries. Library quality control was carried out using an Agilent Bioanalyzer 2100. Expression libraries were sequenced on an Illumina NovaSeq 6000 sequencer.

### Validation experiments confirming the involvement of VKORC1 in NAFLD in HepaRG cells

Single sgRNA transductions were performed as previously described, and HepaRG cells were selected on either BFP expression (for mRNA isolation) or puromycin resistance using (for confocal microscopy. sgRNAs were either targeting *VKORC1*, or were non-targeting sgRNAs as control. Cells were cultured either on standard 6-well plates (mRNA) or in confocal compatible cell culture chambers.

After 10 days of gene-editing, double positive (BFP and mCherry) HepaRG cells were sorted, and total mRNA was isolated using Qiagen kits according to the manufacturer’s instructions. mRNA was reversely transcribed using a High-Capacity cDNA Reverse Transcription kit (Thermofisher). qPCR using TaqMan Master Mix was carried out according to the manufacturer’s instructions on a StepOnePlus Real-Time PCR system from Applied Biosystems, using primers and probes targeting *VKORC1* and *PLIN2* (Thermofisher).

Confocal microscopy was carried out on transduced HepaRG cells selected for puromycin resistance after 10 days of gene-editing. Briefly, the chamber slides were washed and treated with 0.5% BSA and 0.03% Triton-X in PBS to block unspecific binding. Samples were incubated at RT for 1h with 1ug/ml of PLIN2 antibody prepared in 0.1% BSA and 0.03% Triton-X in PBS. After three washes with PBS, samples were incubated with a goat anti-rabbit Alexa Fluor 564 antibody at a 1:5000 dilution at room temperature for 1 hour. Samples were washed with PBS and incubated with 1 µg/ml Bodipy 493/503 (ThermoFisher #D3922) for 30 mins at room temperature, after which mounting media containing DAPI was applied. Cells were imaged on the Leica TCS SP8, using UV light to capture DAPI, and 488 and 552 nm lasers to image lipid droplets using Bodipy staining and PLIN2 staining, respectively.

Image analyst was blinded to the treatments when assessing lipid droplet number, size, area, and PLIN2 staining intensity. Imaging data were analyzed using a custom pipeline in Cell Profiler1 v4.2.5. For each imaging site, background was subtracted using CorrectIlluminationCalculate and CorrectIlluminationApply for each of the channels. Next, nuclei were identified in the DAPI channel using IdentifyPrimaryObjects. BODIPY channel underwent additional thresholding, followed by the identification of lipid vesicles using IdentifyPrimaryObjects and their measurement by MeasureObjectSizeShape. To quantify perilipin staining, the number of pixels above a set threshold were calculated using Threshold and MeasureImageAreaOccupied.

### Transcriptome analysis in human NAFLD

Publicly available data were downloaded from the Gene Expression Omnibus (GEO, GSE130970), where whole genome transcriptomes from 78 patients in differential stages of NAFLD progression are available. Data were downloaded as per GEO instructions, and *VKORC1* expression was assessed in relation to the available metadata on steatosis grade, NAFLD activity score, and inflammation. Expression levels were extracted using R version 4.3.1, and later plotted in GraphPad Prism v.9.

### PheWAS and eQTL analyses in public databases

The global biobank engine was used to explore the lead NAFLD-S SNP (rs9934438) in relation to other cardiometabolic traits (Global Biobank Engine, Stanford, CA (http://gbe.stanford.edu) [12, 2023]). The GTEx database was adapted to explore eQTL effects of rs9934438, (GTEx Portal [https://gtexportal.org/home/] on 12/11/23).

### Statistical analyses

All statistical analyses pertaining the generation of the NAFLD score were carried out in R 3.5.1 using the *pROC* package. Plotting of GWAS results was also carried out using R. Plink v.2 was used for genetic analyses. Single-cell RNA-seq data comparing proliferative and differentiated HepaRG cells were analysed using Seurat, and the ‘FindMarkers’ differential expression analysis function using Wilcoxon tests. Unsupervised clustering based on single-cell transcriptomes was carried out for both differentiated and undifferentiated HepaRG cells. For each cluster, the top 20 upregulated genes in that cluster were input into GSEA pathway enrichment analyses using ‘Hallmark genes’, ‘Gene Ontology’ and ‘Reactome’ pathways. The scMAGeCK package was used to explore differential expression produced by sgRNA perturbations. Differentially expressed genes (comparing targeting vs. non-targeting sgRNAs) from scMAGeCK analyses were input into GSEA pathway enrichment analyses.

Functional experimental data were plotted and analysed using GraphPad Prism v. 9. Student’s T-test, or ANOVA with post-hoc test was applied as appropriate on data passing Shapiro-Wilk’s test for normality, otherwise non-parametric tests were applied.

## Results

### Anthropometric and biochemical data predict NAFLD in UKB

We explored existing data on liver fat percentage obtained from abdominal MRI in UKB. NAFLD was defined as a liver fat percentage >5.5%, resulting in 2,544 NAFLD cases and 10,168 controls. Anthropometric and biochemical variables related to NAFLD and cardiometabolic traits were interrogated for their ability to predict NAFLD defined as above using multivariate regression models. The variables included in the regression model can be found in ***Table 1***. Predictors of NAFLD were selected to create a NAFLD score using ***Equation 1***.

### NAFLD score improves NAFLD approximation

The power to approximate NAFLD using the generated NAFLD score (NAFLD-S) was assessed using a receiver operating characteristic (ROC) curve, and the area under the curve was compared between NAFLD-S, FLI and several individual anthropometric and biochemical variables. Our results reveal that NAFLD-S improves approximation of NAFLD status compared to FLI. Further, NAFLD-S outperformed all individual anthropometric and biochemical variables on which the NAFLD-S was based, ***Figure 1A***.

**Figure 1.**
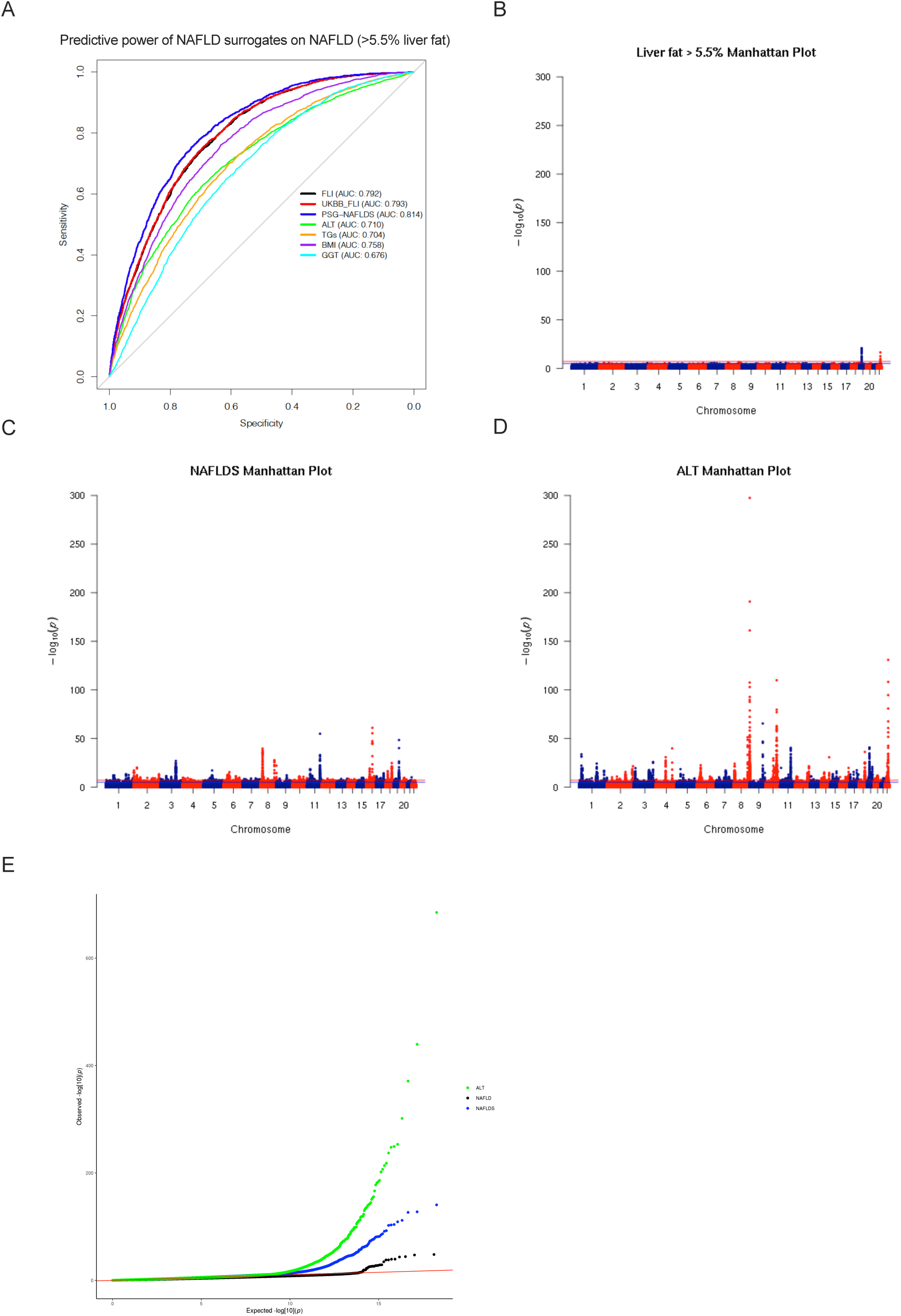
Human molecular genetic analyses in the UK Biobank. Non- or moderately drinking European ancestry British participants were selected for the analyses. **A)** ROC curve showing the predictive power of NAFLD-S and individual biochemical and anthropometric variables on NAFLD status as defined by liver fat > 5.5% in UK biobank. **B)** Manhattan plot for the genome-wide association study on NAFLD defined as >5.5% liver fat in UK biobank. **C)** Genome-wide association study on NAFLD score in UK biobank, visualized using a Manhattan plot. **D)** ALT associations from the genome-wide association study in UK biobank, visualized by a Manhattan plot. **E)** Q-Q plot for the GWA studies on ALT, NAFLD, and NAFLD score, plotted together to visualize the differences in significance obtained. Y-axes in Manhattan plots are scaled for comparison between the three association studies.

The NAFLD-S was calculated for a subset of non- and moderate drinkers in the UKB, and a GWAS was carried out on NAFLD-S as a continuous variable. In parallel, GWAS were carried out for liver fat percentage (MRI_UKB), and ALT (qnormALT_UKB) in the same subset of UKB. Our results show numerous associations to liver fat percentage, NAFLD-S, and ALT (***Figure 1B-D***). There is a sizable overlap in loci that are associated with NAFLDNAFLD-S and liver fat percentage. For example, the *PNPLA3* locus is detected in GWAS of NAFLD-S, ALT, and liver fat percentage. However, since ALT and NAFLD-S use a larger portion of the UKB, there is a substantially larger number of associations for ALT and NAFLD-S, compared to liver fat percentage. Another effect of the larger sample size used in the association studies for ALT and NAFLD-S is the typically smaller p-value for these associations, which is visualized by scaled y-axes on Manhattan plots, and the three association studies plotted in the same Q-Q plot, ***Figure 1E***. Summary statistics for significant associations can be found in ***Supplementary Tables 1-3***.

To aid in inferring causality and to prioritize genes for functional follow-up, we assessed GWAS SNPs associated with liver fat percentage and common surrogates through genetic colocalization to eQTL and sQTLs from GTEx (v8) using our custom pipeline (15). For this analysis, we used our NAFLD-S as well as a previously published score, recently published data on liver enzymes, chronically elevated ALT and MRI/ML ***Figure 2A*** to explore the overlap between different approaches to detect genetic associations and causal genes for NAFLD (23–25). Genes demonstrating a significant colocalization to the liver tissue in the GTEx data base were prioritized.

**Figure 2.**
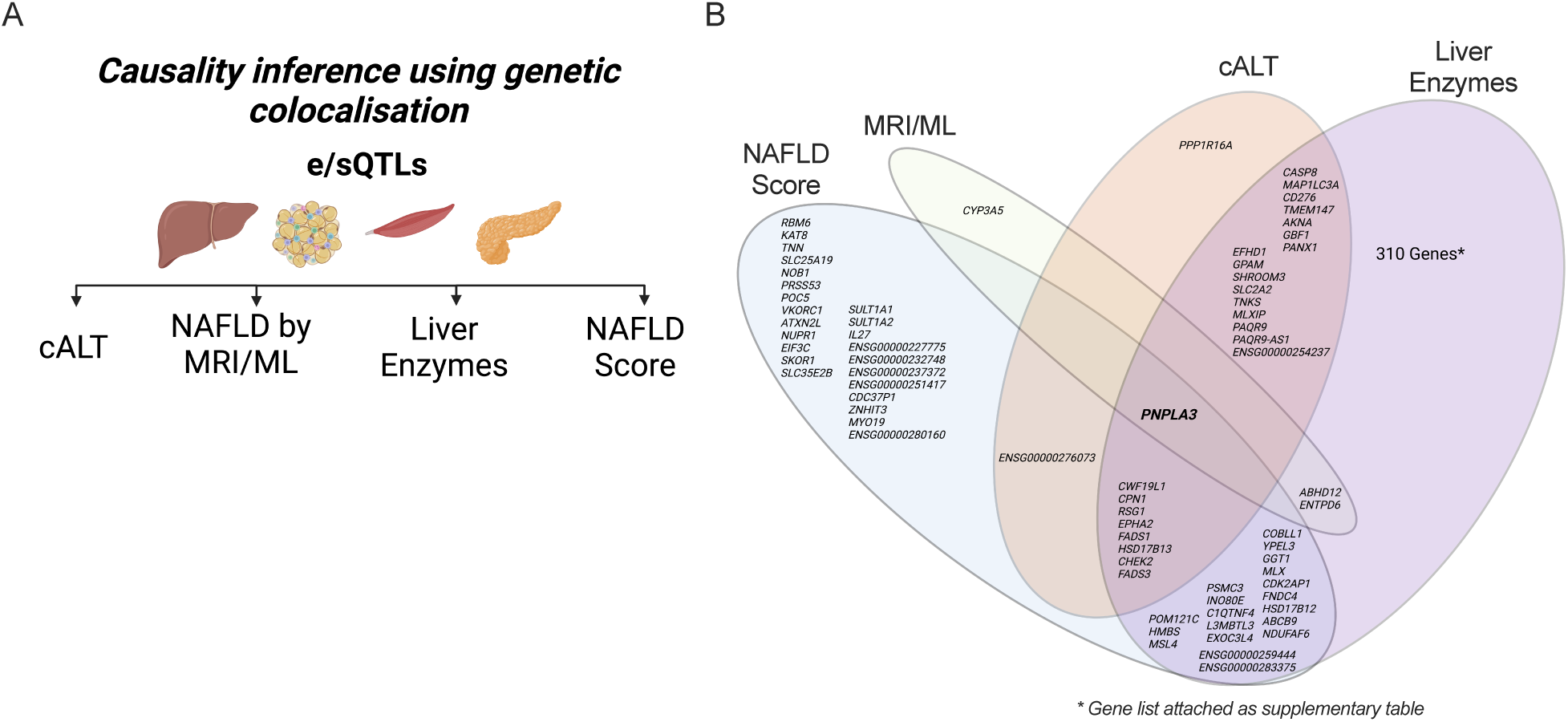
Colocalization study of NAFLD-S associated SNPs in the UK Biobank. **A)** Strategy for genetic colocalization studies to infer causality of novel putative NAFLD genes found from GWAS with metabolically active tissues in the GTEx (v8) database. Liver enzymes include ALT, ALP, GGT and qnormALT_UKB, MRI/ML MRI_UKB and machine-learning MRI, NAFLD-S out novel NAFLD score and the NAFLD score from Miao et al. **B)** Overlap of genes with a significant liver eQTL/sQTL colocalization. GSEA gene set enrichment analysis of colocalized genes can be found in ***Table 2***. Full list of colocalizations can be found in ***Supplementary Table 5***.

Due to the relative paucity of GWAS data for liver fat percentage, only four colocalizations were found for our MRI/ML in liver: *PNPLA3* (which is also shared with all NAFLD surrogate markers), *CYP3A5*, *ABHD12*, and *ENTPD6*. In contrast, we observed numerous colocalizations originating from GWAS of NAFLD surrogates, with sizable overlap between the different surrogates, ***Figure 2B*** and ***Supplementary Table 4***. Numerous other genes that have previously been suggested to influence NAFLD were also found to show significant colocalization; e.g., in or near *GPAM* (ALT), *AKNA* (ALT and NAFLD-S), and the *TNKS*/*PPP1R3B* (ALT). *VKORC1*, a gene previously associated with triglyceride levels and body fat distribution, colocalizes with NAFLD-S (26, 27). We then used colocalized NAFLD-S genes as input in a GSEA pathway enrichment analysis, and show that these genes are enriched in processes related to lipid homeostasis, steroid and lipid metabolism, and sterol homeostasis, ***Table 2***. Colocalized ALT genes, however, are primarily enriched in processes pertaining to organelle organization, small molecule metabolic processes, response to stress, and lipid metabolism, ***Supplementary Table 5***. Importantly, our NAFLD-S outperforms ALT in approximating liver fat >5.5% and variants associated with NAFLD-S more often co-localize with lipid metabolism-related eQTLs in liver (n=XXXX genes) than variants associated with ALT (n=XXX genes). This provides a rationale for using composite surrogate variables for GWAS of NAFLD as these may capture more of the biology of the disease and provide better insight in the natural history of NAFLD than single biochemical surrogates.

Collectively, these data not only suggest that creating composite surrogate markers for NAFLD may be used to identify putative NAFLD genes when there is a paucity of gold standard MRI data, but also that there may be biological differences driving the different associations with surrogate phenotypes, which has implications for the pathogenesis of NAFLD.

### Establishing a HepaRG cell line suitable for genome editing

To do functional follow up studies following gene knockdown experiments, we genetically engineered HepaRG cells to stably express dCas9-KRAB, which allows for CRISPRi. The introduction pHR-SFFV-KRAB-dCas9-P2A-mCherry into HepaRG cells allows for transcriptional interference of genes targeted by sgRNAs by KRAB. The resulting cell line (dCas9-KRAB-HepaRG) was used to characterize putative NAFLD genes. HepaRG cells underwent single-cell RNA sequencing (scRNA-seq) to characterize the model system and ensure that the introduction of dCas9-KRAB does not alter the function and the ability to differentiate of HepaRG cells. dCas9-KRAB-HepaRG cell line was efficiently differentiated using established protocols, ***Figure 3A***, did not differ at the transcriptome level, assessed by scRNA-Seq, regardless of dCas9-KRAB integration, and assumed a hepatocyte-like phenotype upon treatment with a differentiation media for two weeks ***Supplementary Figure 1***.

**Figure 3.**
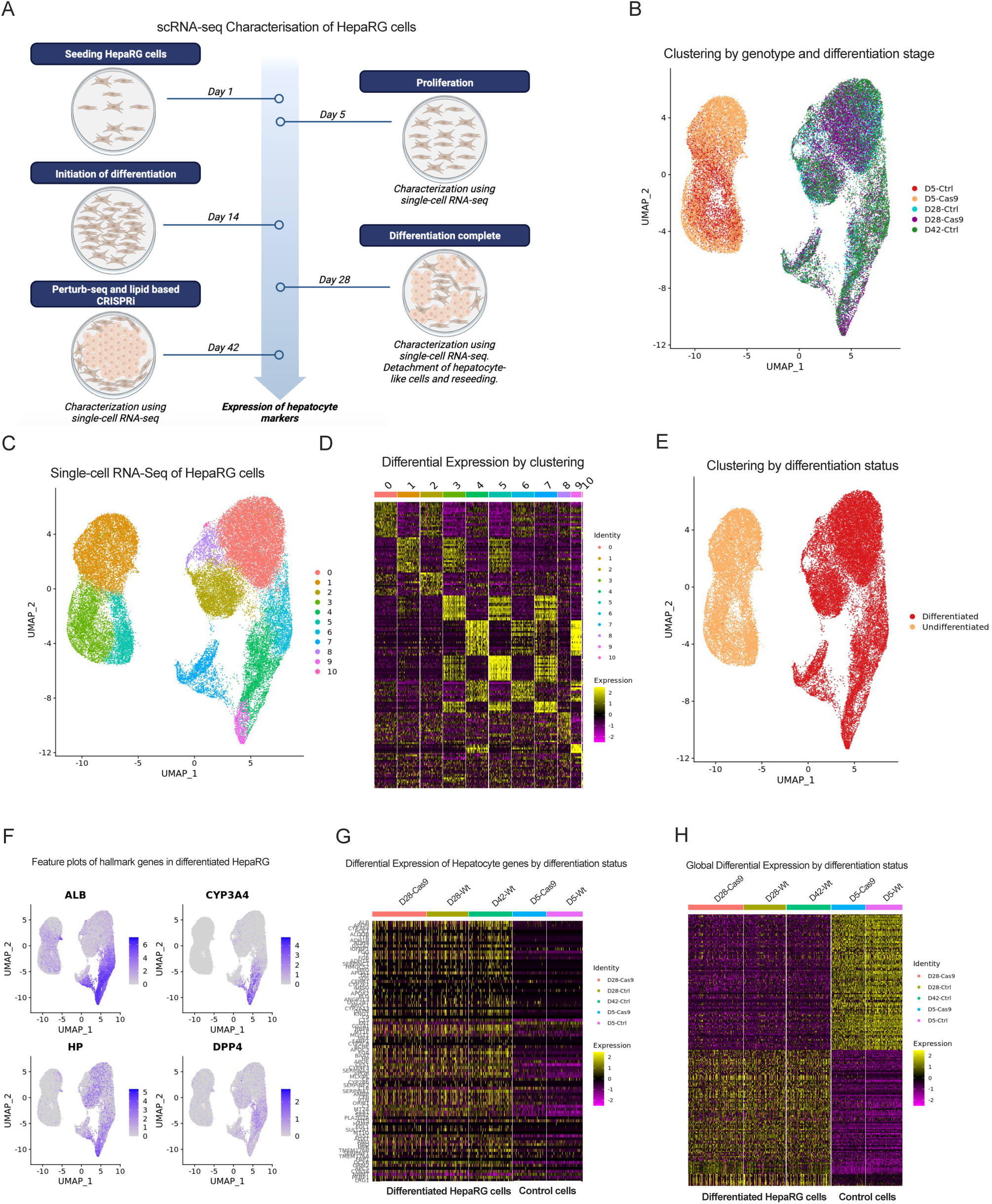
Characterisation of a HepaRG model system that is genetically engineered to allow for CRISPRi gene-editing. **A)** Description of HepaRG culturing, indicating at which point scRNA-seq was used to characterize the model system. **B)** Clustering by both genotype and differentiation stage (temporal analysis along the differentiation axis). Data demonstrate that cells efficiently differentiate regardless of genotype (dCas9-KRAB integration) and that cells remain in their differentiated phenotype two weeks after differentiation is complete. This allows for gene-editing after complete HepaRG differentiation. **C)** Clustering of scRNA-seq data, where proliferative and differentiated cells are plotted together, irrespective of genotype (dCas9-KRAB integration). Data show 11 different clusters divided over two distinct populations of cells. **D)** Differential gene expression analyses based on clustering in Figure 3B. Clusters 1, 3 and 5 belong to undifferentiated cells, whereas the remaining clusters belong to the differentiated HepaRG cells; genes involved in drug metabolizing pathways, lipid metabolism, hemostasis and albumin were significantly upregulated in differentiated cells, particularly in clusters 0, 2, 4, 6 and 9. Lists for differentially expressed genes can be found in ***Supplementary Table 6***. **E)** Clustering by differentiation status irrespective of genotype. Data demonstrate a perfect clustering of HepaRG cells by their differentiation status. **F)** Expression of hepatocyte hallmark genes *ALB*, *CYP3A5*, *HP-1* and *DPP4*. Data show an upregulation of these genes upon differentiation. **G)** Differential expression analyses of genes suggested to define hepatocytes from ‘*the human liver atlas*’. Data show that the transcriptional program thought to define hepatocytes is enhanced upon differentiation. **H)** Global differential expression analyses by differentiation status. Genes upregulated by differentiation are enriched in processes related to small molecule and lipid metabolic processes, mitochondrial processes and electron transport chain, ***Supplementary Table 10***. Complete lists of differentially expressed genes can be found in ***Supplementary Table 7***.

Standard scRNA-seq quality control steps were taken, and revealed that, upon differentiation, HepaRG cells increased their expression of mitochondrial genes, while the overall number of genes expressed at detectable levels was marginally decreased, ***Supplementary Figure 2***. The upregulation of mitochondrial genes was not surprising as upon differentiation, HepaRG cells have been documented to increase their metabolism while suppressing proliferation. Clustering of single-cells showed that there are no significant differences between wild-type (Wt) and genetically engineered dCas9-KRAB-HepaRG cells, ***Figure 3B***, and thus, all cells were analyzed jointly.

Clustering with regard to single-cell transcriptomes of Wt and dCas9-KRAB HepaRG cells revealed 11 distinct clusters; clusters 1, 3 and 5 belong to undifferentiated cells, whereas the remaining clusters belong to differentiated HepaRG cells. Genes involved in the cell cycle, G2M checkpoint, EMT and cell division are all more highly expressed in the undifferentiated clusters. In contrast, genes involved in drug metabolizing pathways, lipid metabolism, hemostasis and albumin are significantly upregulated in differentiated cells, particularly in clusters 0, 2, 4, 6 and 9, ***Figure 3C-D, Supplementary Table 6*.** It is expected that numerous cells undergo apoptosis during the differentiation process. In line with this, cells within clusters 8 and 10 express genes involved in apoptosis, p53 and programmed cell death, ***Figure 3D, Supplementary Table 6*.** Cells within cluster 7 seem to consist of a population of cells that may not be fully differentiated, as they highly express some hepatocyte and proliferative markers, ***Figure 3D*** and ***Supplementary Table 6***. In summary, HepaRG cells are efficiently differentiated to a hepatocyte-like phenotype as the transcriptome of the cells belonging to clusters 4, 6, 7, 9 (>50% of cells) indicate a shift consistent with hepatocyte biology and function.

Differentiated and proliferative HepaRG cells cluster separately, as shown in **Figure 3E**. We compared the differentiated and undifferentiated cell populations based on a list of hepatocyte markers and the human liver atlas (28), regardless of their dCas9-KRAB status. Differentiated cells demonstrate an increased expression of hallmark hepatocyte genes including *ALB*, *CYP3A4*, *HP*, and *DPP4*, ***Figure 3F***. The expression of a list of hepatocyte genes involved in drug and lipid metabolism were also increased compared to undifferentiated cells, ***Figure 3G***. Next, we analyzed global differential gene expression. Gene set enrichment analysis revealed that differentiated HepaRG cells increase their expression of genes involved in metabolic processes; both in lipid metabolism and the genes within drug metabolism, ***Figure 3H, Supplementary Figure 3A-B***, and ***Supplementary Table 7***.

### CRISPRi screen and Perturb-seq implicates putative causal genes in NAFLD

We created a combinatory lipid accumulation-based CRISPRi and Perturb-seq screen in the dCas9-KRAB-HepaRG cell system to investigate putative NAFLD genes. We optimized the lipid accumulation-based CRISPRi system by knocking down the lipid droplet associated protein *PLIN2* to markedly reduce lipid accumulation. dCas9-KRAB-HepaRG cells were transduced with three sgRNAs targeting *PLIN2* along with a non-targeting sgRNA as a control, loaded with 400uM oleic acid for 24hrs. Next, neutral lipids were stained using Bodipy. Lipid loading was significantly increased after 24hrs of oleic acid treatment, and the efficiency of the sgRNAs was confirmed, ***Figure 4A-C***. mCherry/BFP^+/+^ HepaRG cells were sorted with regard to lipid content after 10 days of gene-editing, and gDNA was isolated in the most and least lipid-laden cells (the 20^th^ percentile in either tail), ***Figure 4D-E***. Sequencing of gDNA from the most and least of lipid loaded HepaRG cells, as measured by Bodypi staining, revealed a significant enrichment of *PLIN2* sgRNAs in the least lipid-laden cells, indicating that *PLIN2* knockdown indeed impairs lipid accumulation, ***Figure 4F-G***. This experiment served as a proof-of-principle for our CRISPRi screen, which included sgRNAs targeting a small selection of putative NAFLD genes, selected based on our human molecular genetic analyses.

**Figure 4.**
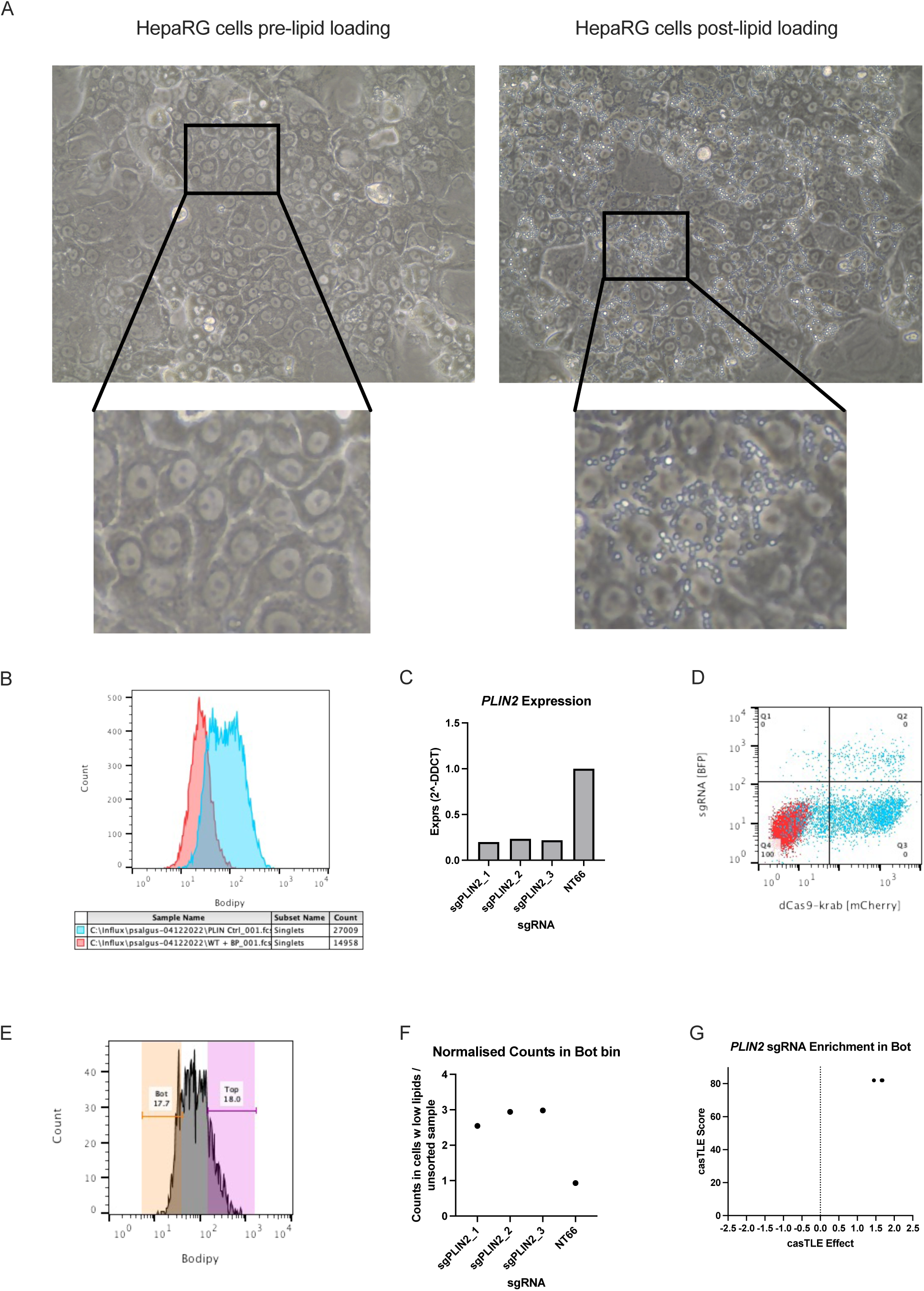
Establishment of, and control experiments in a HepaRG cell CRISPRi gene-editing model system with lipid accumulation as readout. **A)** Micrographs showing that lipid loading using 400 µM of oleic acid results in significant formation of large lipid droplets. **B)** Lipid loaded HepaRG cells were stained with 1 µg/ml Bodipy and analysed using flow cytometry. Data show that lipid loading (Blue histogram) increases the content of neutral lipids within the HepaRG cell compared to non-loaded control cells (Red histogram). **C)** *PLIN2* was knocked down as a proof-of-principle experiment. *PLIN2* expression was efficiently silenced in our dCas9-KRAB expressing HepaRG cells, and sgRNAs from the v2 Weissman library. **D)** Representative gates for sorting gene-edited HepaRG cells (blue), and an untransduced control, negative for both mCherry and BFP (red). The Q2 gate contains the gene-edited cells, which express dCas9-KRAB, and have been efficiently transduced with sgRNAs. **E)** Gene-edited HepaRG cells from Q2 were sorted based on their Bodipy content; approximately the top and bottom 18% of cells were sorted, and gDNA was prepared from both extreme populations. gDNA was then sequenced using NGS. **F-G)** By assessing the enrichment of *PLIN2* sgRNAs in the cell population with the least intracellular lipids, we find a 2-3 times enrichment of *PLIN2* sgRNAs compared to non-targeting sgRNAs. As one would expect, these data demonstrate hampered lipid accumulation in HeapRG cells that do not express *PLIN2*. The casTLE pipeline was also piloted for this purpose, and analysed data recapitulates simple sgRNA counting and fold change calculations in that we show a 2 times enrichment of PLIN2 sgRNAs in the least lipid laden cells compared to what one would expect by chance. Data show that the model system can provide useful information on the effect of genes on lipid accumulation in HepaRG cells, and show that appropriate analysis methods are employed.

Eleven known and putative NAFLD genes were selected for tandem CRISPRi and Perturb-seq to explore their role in NAFLDNAFLD development, as measured by HepaRG lipid accumulation and single-cell transcriptional changes. The genes were selected based on 1) their robustness of association to NAFLD NAFLD (amount of evidence if known NAFLD gene), 2) emerging evidence for an association without functional validation, and 3) new association with NAFLD-S that also demonstrates association to other cardiometabolic traits. The genes can be found in ***Table 2***. Next, sgRNAs directed towards selected NAFLD genes were transduced in differentiated dCas9-expressing HepaRG cells for a tandem CRISPRi and Perturb-seq experiment ***Figure 5A***.

**Figure 5.**
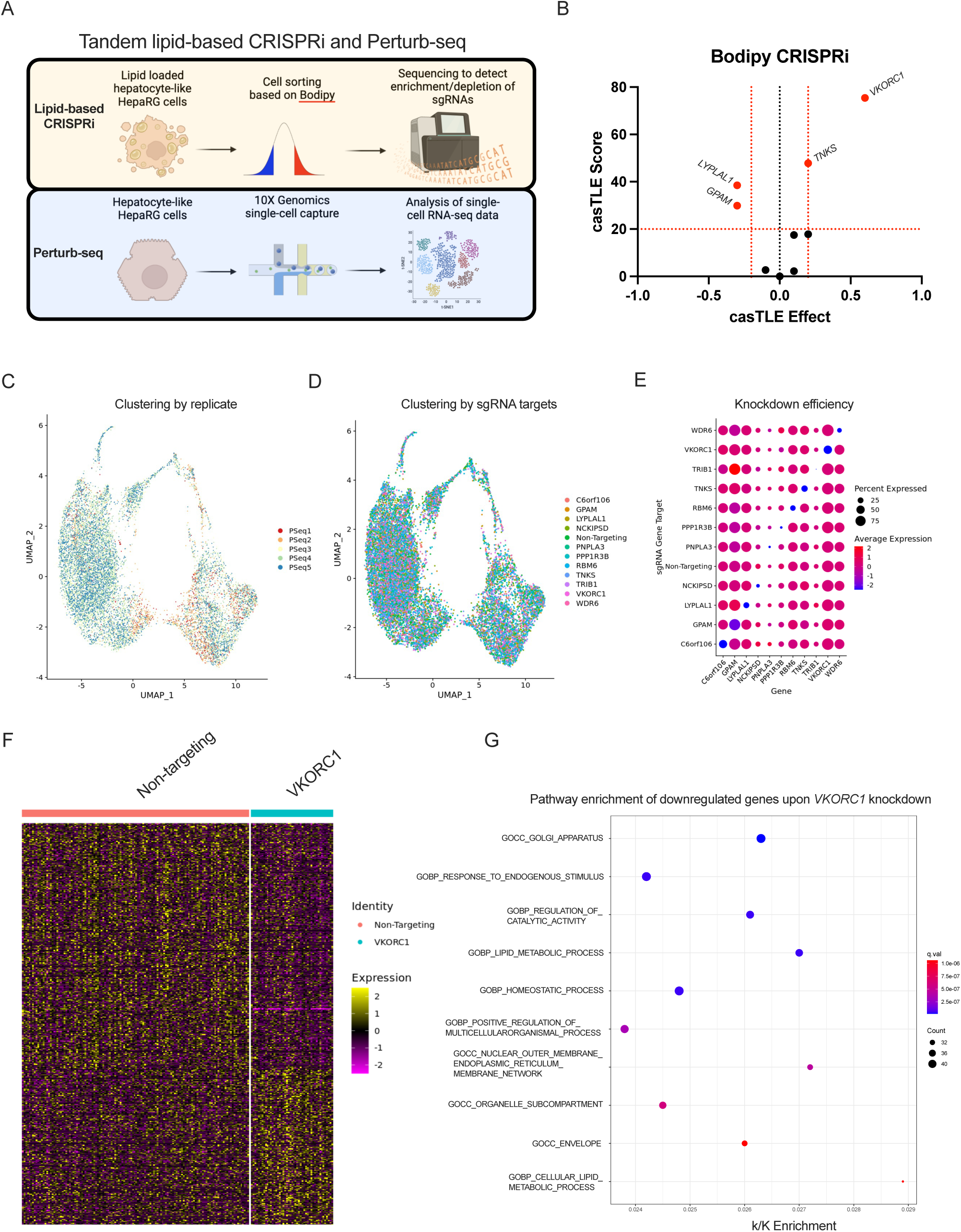
Tandem lipid-based CRISPRi and Perturb-seq in HepaRG cells to explore the involvement of genes, suggested from human molecular genetics, in NAFLD pathogenesis. **A)** Experimental outline of tandem CRISPRi and Perturb-seq in HepaRG cells. HepaRG cells were harvested on day 42 of culturing, as per the protocol described in Figure 3A. **B)** Volcano plot, following sequencing of gDNA in the most and least lipid laden HepaRG cells, where casTLE effect and score are plotted against each other. Data demonstrate that knockdown of *VKORC1* and *TNKS* results in less intracellular lipids. Conversely, knockdown of genes *GPAM* and *LYPLAL1* increases intracellular lipids. **C-D)** Perturb-seq is performed in parallel to our lipid accumulation-based CRISPRi to explore the transcriptomic profiles resulting from a gene knockdown. No major changes in clustering of gene-edited cells by replicate and sgRNA identity is observed. Data show that replicates are very similar, and sgRNAs have modest effects on the transcriptome that causes the cells to cluster separately. **E)** Dotplot visualizing that the knockdown of sgRNAs targeting the selected genes is efficient and specific as demonstrated by the blue dots along the diagonal. **F-G)** Differential gene expression analyses upon *VKORC1* knockdown are carried out using the scMaGeCK R-package, and differentially expressed genes are plotted in a representative heatmap. Results reveal that *VKORC1* knockdown changes the transcriptional landscape, and reduces the gene expression of genes involved in lipid metabolism, Golgi and ER, as well as homeostatic processes. Complete results of differentially expressed genes for all perturbations can be found in ***Supplementary Figure 4***, and ***Supplementary Table 8***.

Cells were loaded with oleic acid and then sorted based on mCherry/BFP^+/+^, and their lipid accumulation was measured by Bodipy staining. Sequencing of gDNA from either extreme population with regard to lipid accumulation (∼ top/bottom 15%) revealed that *VKORC1* and *TNKS* sgRNAs are enriched in the bottom population, whereas *GPAM* and *LYPLAL1* sgRNAs are enriched in the top population. This suggests that *VKORC1* and *TNKS* knockdown reduces lipid accumulation, whereas *GPAM* and *LYPLAL1* knockdown increases lipid accumulation, ***Figure 5B***.

In parallel to the lipid accumulation-based CRISPRi screen, we produced single-cell transcriptomes from all perturbations. The experiment was performed in a total of five 10X Genomics single-cell captures, from two biological replicates. Single-cell transcriptomes were analyzed using Seurat, and the general quality control data is visualized in ***Supplementary Figure 3A***. While there was no clustering by replicate or sgRNA identity, perturbations produced by the sgRNAs are consistently efficient and specific as only the intended target gene is significantly knocked down, ***Figure 5C-E***. *VKORC1* knockdown produced the most striking transcriptional changes, and will be discussed in detail below. *GPAM* knockdown resulted in a downregulation of genes enriched in oxidative phosphorylation and RNA transcription pathways, while the upregulated genes were enriched in pathways pertaining cellular stress, glycolysis, apoptosis and cell cycle, ***Supplementary Table 8***. *LYPLAL1* knockdown resulted in the downregulation of genes involved in interferon-response, adipogenesis, and oxidative phosphorylation, amongst others. Genes upregulated by *LYPLAL1* knockdown are enriched in metabolism of heme and blood vessel formation, ***Supplementary Table 8***. We found pathway enrichments in adipogenesis, HDL, and chylomicron metabolism, and estrogen response among genes downregulated upon *TNKS* knockdown. Differential gene expression for all perturbations are visualized as heatmaps in ***Supplementary Figure 4***, and ***Supplementary Table 8***.

While we decided to focus on the target gene *VKORC1* – because of its novelty and significant impact on lipid accumulation – we validate one gene (*GPAM*) influencing lipid accumulation in the opposite direction to *VKORC1*. We knocked down *GPAM* using single sgRNA transductions, and recapitulated the findings from the lipid accumulation-based CRISPRi screen where *GPAM* knockdown results in an increase in lipid accumulation, ***Supplementary Figure 5A-D***.

### VKORC1 is involved in the development and progression of hepatosteatosis

Differential gene expression as a result of *VKORC1* knockdown was investigated over the two replicates of Perturb-seq experiments using the scMAGeCK package in R. All differentially expressed genes from the two replicates were investigated for gene set enrichment, and results show that genes enriched in lipid metabolic pathways are downregulated upon *VKORC1* knockdown, ***Figure 5F-G*** and ***Supplementary Tables 9-11***. Further, agnostic differential gene expression analyses demonstrate that *VKORC1* knockdown alters the expression of a set of genes related to liver lipid metabolism and insulin resistance, ***Supplementary Figure 5E***. Specifically, under *VKORC1* knockdown conditions there is a trend for reduced expression in cells of genes involved in lipoprotein production and secretion (*DGAT1*, *DGAT2*, *APOB*, *APOC1*, and *MTTP*), and of the lipid accumulation marker *PLIN2*. Our scRNA-seq data reinforces the notion that *VKORC1* may influence lipid accumulation and *PLIN2* expression since there is a correlation between *PLIN2* and *VKORC1* expression in cells transduced with non-targeting sgRNAs in our Perturb-seq experiments, ***Supplementary Figure 5F-G***.

Further, metabolic perturbations in another hepatocyte model system, HepG2 cells, revealed that *VKORC1* expression is reduced by glucose, and shows a trend towards downregulation by atorvastatin, ***Supplementary Figure 5H***.

We construct a protein-protein interaction network using BioGRID to explore what proteins might interact with VKORC1. Analyses reveal that there is a physical interaction with apolipoproteins, which reinforces the notion that VKORC1 may have a previously unexplored role in liver lipid metabolism, ***Supplementary Figure 6A***. We investigate gene set enrichment of all VKORC1 interactors, and enrichments were found in processes pertaining lipid homeostasis, oxidoreductase activity, lipid metabolic processes, and sterol homeostasis, ***Supplementary Table 11***.

We next performed knockdown experiments of *VKORC1* in HepaRG cells using single sgRNA transductions to confirm our observations from single-cell CRISPRi screens, with a non-targeting sgRNA as control. The knockdown was confirmed using RT-qPCR against *VKORC1*, **Figure 6A**. We recapitulated the *in vitro* phenotype observed in the single-cell CRISPRi screens, where the reduction of *VKORC1* expression brought about a reduction in *PLIN2* expression, accompanied by a reduction in lipid accumulation as measured by Bodipy using flow cytometry and confocal microscopy, ***Figure 6B-E***.

**Figure 6.**
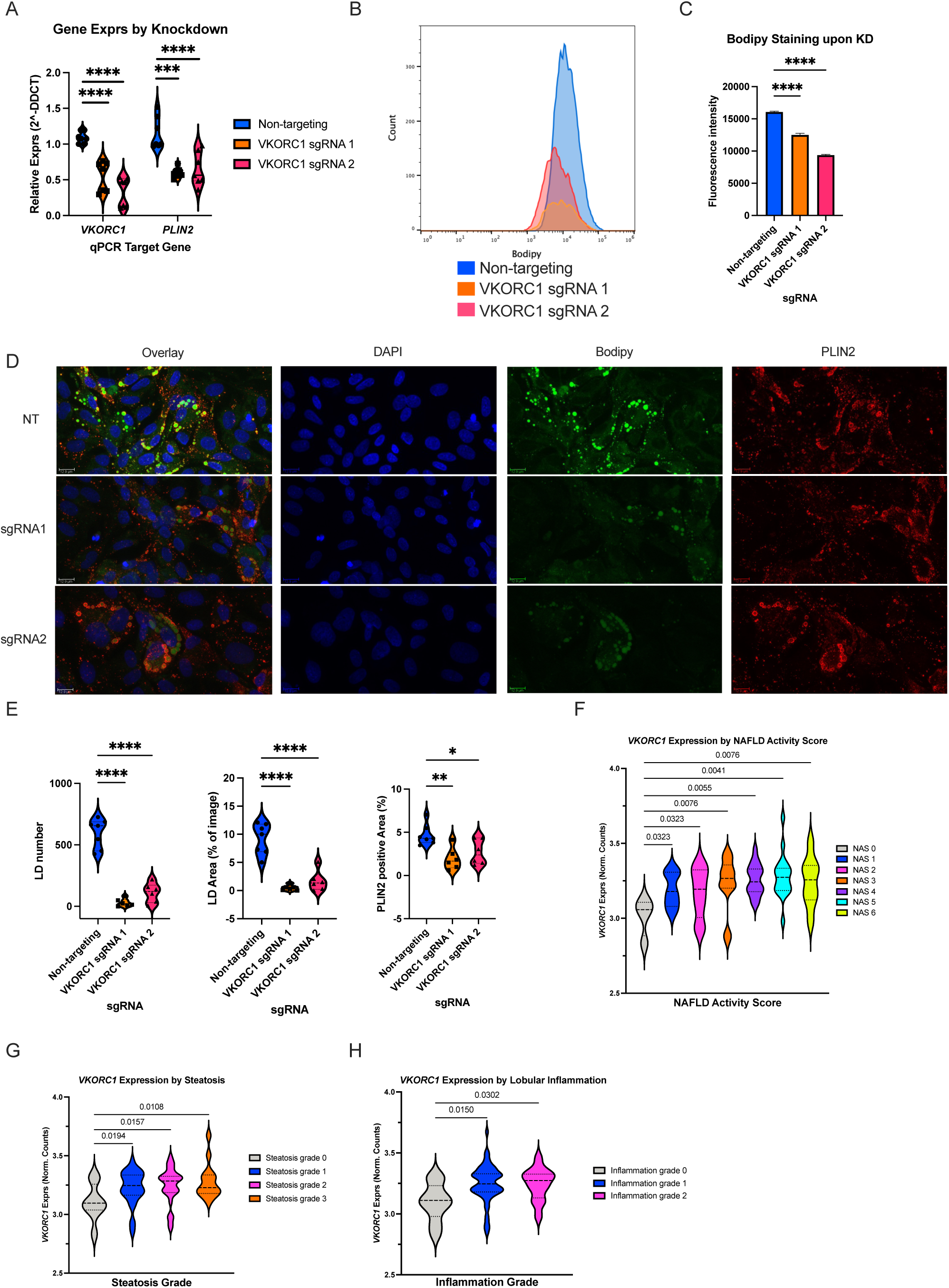
Validation experiments of *VKORC1* knockdown in differentiated HepaRG cells, and the relationship between *VKORC1* transcript and human disease. **A)** Single sgRNA knockdown of *VKORC1* in differentiated HepaRG cells results in a significant knockdown of the *VKORC1* transcript as measured by qPCR. Concomittant with *VKORC1* knockdown, we demonstrate a significant downregulation of the *PLIN2* transcript. **B-C)** *VKORC1* knockdown results in reduction of intracellular neutral lipids by Bodipy staining and flow cytometric analysis. **D-E)** Confocal microscopy of HepaRG cells upon *VKORC1* knockdown shows a significant reduction in Bodipy neutral lipid staining; lipid droplet number, lipid droplet area, as well as PLIN2 positive area. **F-H)** By exploring *VKORC1* expression levels in different stages of human disease we demonstrate an upregulation of the *VKORC1* transcript in livers of a higher degree of NAFLD activity score, steatsis, and inflammation. *N for experimental data is 6-7 replicates, Ordinary one-way ANOVA was performed to compare the non-targeting sgRNA with the VKORC1 targeting sgRNAs*. *Total n for human liver samples is 78*. *\* p<0.05, ** p<0.01, ***p<0.001, ****p<0.0001*.

To better understand the role of *VKORC1* expression in human NAFLD, we investigated publicly available data on *VKORC1* transcript levels in a cohort of 78 human livers encompassing the entire spectrum of NAFLD. Our analyses show a positive association of NAFLD activity score, steatosis, and inflammation with *VKORC1* expression, ***Figure 6F-H***. These data suggest that *VKORC1* is involved in the initiation of NAFLD, however, *VKORC1* does not seem to the primary driver of the progression of disease as transcript levels only increase over the lowest grade of disease, and not as grades of disease progress.

We explored the co-expression patterns of *VKORC1* and transcripts of a selection of genes involved in lipid metabolism and fibrosis that are thought to drive disease progression in healthy human liver, the ASAP study. Co-expression patterns revealed that *VKORC1* expression correlates with the expression of genes involved in uptake of lipids, as well as in intracellular fatty acid and triglyceride synthesis. Further, *VKORC1* mRNA levels are correlated with transcript levels of collagen and TGFβ, which are genes known to promote fibrosis, ***Supplementary Figure 7A***. *VKORC1* was negatively correlated with genes involved in the mobilization of lipids from hepatocytes; *MTTP* and *SREBF1*, suggesting that *VKORC1* expression promotes the intracellular accumulation of lipids in human liver.

The *in vitro* NAFLD phenotype is also recapitulated in mice fed a high fat diet for 30 weeks, known to induce NAFLD, where both *Vkorc1* and *Plin2* expression is concomitantly increased in animals on high fat diet, ***Supplementary Figure 6B***.

Further exploration of genetic data, and PheWAS revealed a large LD-block in the NAFLD-S-associated *VKORC1* locus, and the A allele is associated with a lower *VKORC1* expression in liver (GTEx v8 database), reinforcing the observed relationship between *VKORC1* and an *in vitro* NAFLD phenotype, as well as the phenotype obtained from *in vivo* models of disease, ***Supplementary Figure 8A-B***. All the SNPs in the *VKORC1* locus that contribute to the NAFLD-S association, and colocalization are associated with the expected (protective) effect with regard to cardiometabolic risk factors/biomarkers, ***Supplementary Figure 8C***. The NAFLD-S reducing A allele of lead SNP rs9934438 is also associated with reduced lower hip and waist circumference, BMI, and numerous fat mass phenotypes as well as a protective association with numerous biomarkers of cardiometabolic disease; including lower plasma triglycerides, ApoB, HbA1c, and higher HDL and ApoA, ***Supplementary Figure 8D***.

In summary, we have demonstrated the usefulness of using NAFLD-S as a surrogate marker for NAFLD, prioritized candidate NAFLD genes from past and the present study using a custom genetic colocalization analysis for functional follow-up. After assigning putative causal genes for functional follow-up coming from GWAS for different NAFLD surrogates, we performed a functional CRISPRi screen for lipid accumulation and Perturb-seq transcriptional analysis at a single cell level, which constitutes a functional genomic framework and allows for interrogation of putative NAFLD genes at scale. By using our functional genomics framework; originating from human genetics, moving to functional *in vitro* studies, and later to murine and human disease, we propose that *VKORC1* is implicated in the pathogenesis of NAFLD. Our data suggest that *VKORC1* expression is associated with the increase in intracellular accumulation of lipids, and thereby drive the intiation of NAFLD development. Investigations can now be expanded to interrogate a large selection of putative causal NAFLD genes to further determine the molecular landscape of disease development and progression.

## Discussion

In the present study we generate a NAFLD-S that outperforms single variable surrogates when validated against ‘ground truth’ NAFLD as defined by >5.5% liver fat obtained from proton density fat fraction from MRI images in the UKB. By using a new surrogate marker of NAFLD for our GWAS, together with colocalization analyses of previous GWAS, we expand the knowledge on the genetic susceptibility to NAFLD. We create a functional genomic framework to validate putative NAFLD genes and explore the role of a subset of genes on hepatocyte lipid accumulation, single-cell transcriptomes, and murine and human disease.

GWAS have been useful in the identification of common susceptibility variants for various cardiometabolic traits. However, GWAS for NAFLD have remained small and underpowered, and therefore, surrogate markers of NAFLD have been extensively used, all with their strengths and drawbacks. The FLI does not seem to outperform waist circumference in predicting NAFLD (4), NAFLD may be present without ALT elevation (29,30), and high ALT levels could reflect a myriad of liver insults. Thus, FLI and ALT may constitute poor surrogates for NAFLD. Our NAFLD-S might not only reflect metabolic liver disease, but also an insulin resistance phenotype as the score takes into account waist circumference, BMI, HbA1c and triglyceride levels, which are all also associated with insulin resistance. By integrating anthropometric and biochemical data into a single score, we sought to capture the global etiology of NAFLD given its correlation with dyslipidemia, type II diabetes, and obesity. We outperform ALT levels in predicting liver fat in the UKB, however, mitigation of the drawbacks of using ALT measurements as surrogate for NAFLD could be achieved by using chronic ALT elevation, which has been described elsewhere. (24) We find several overlaps in the genetic colocalizations between several NAFLD surrogates, which suggests that these surrogates capture a common part of the disease etiology, but also may reflect different aspects of the natural history of NAFLD.

We created a HepaRG cell model system suitable for large-scale CRISPRi screening and Perturb-seq to explore putative NAFLD genes. We selected a group of both previously associated (*PNPLA3*, *TNKS*/*PPP1R3B*, *GPAM*, *LYPLAL1*, *TRIB1*) and novel or less established (*WDR6*, *VKORC1*, *RBM6*, *NCKIPSD*, *C6orf106*) NAFLD candidate genes to establish a functional genetic framework to interrogate new potential disease genes at scale. We screened the selected genes for their influence on hepatocyte lipid accumulation in our *in vitro* gene-editing system, and generated single-cell transcriptomes for these CRISPR-based knockdown perturbations. Our data suggest the involvement of *VKORC1* and *TNKS* (increased lipid content), *LYPLAL1* and *GPAM* (decreased lipid content) on lipid accumulation in hepatocytes, which we further validated for *VKORC1* and *GPAM*. The latter is a well-established NAFLD locus, and its knockdown resulted in increased lipid accumulation in HepaRG cells. In contrast, *VKORC1* knockdown resulted in less lipid accumulation, possibly mediated by lower *PLIN2* levels. The knockdown of *VKORC1* also resulted in the perturbation of the transcriptional landscape of lipid metabolism, and insulin resistance genes like *PLIN2*, *PNPLA2*, *G6PC*, and *INSR* were dysregulated. Finally, we explored the mRNA expression of *VKORC1* in a murine model and human disease, where *VKORC1* expression consistently was increased upon high-fat diet. We strengthen the notion that low *VKORC1* expression may be protective of disease development, by exploring the expression levels in relation to the degree of steatosis, NAFLD activity score, and inflammation. By using human molecular genetics, we demonstrated that the NAFLD-S lowering SNP rs9934438 also improves other anthropometric and biochemical cardiometabolic traits, while lowering the expression of the *VKORC1* transcript. Collectively, this suggests a protective role of low *VKORC1* expression in NAFLD.

VKORC1 is known to reduce vitamin K to its active form, which promotes the formation of functional clotting factors from pro-clotting factors. This process is inhibited by warfarin and ultimately results in reduced activation of coagulation factors IX, VII and prothrombin, which is how warfarin exerts its antithrombotic effects. (31) Some studies have indeed described an association between thrombotic risk factors and extent of fibrosis in NAFLD. (32) Similarly, researchers have found elevated, and increased activity of coagulation factors in NAFLD. (33) Likewise, it has been observed that there is a higher-than-expected prevalence of NAFLD in patients suffering from idiopathic venous thromboembolism. (34) However, to the best of our knowledge, we provide the first data implicating *VKORC1* in the hepatocyte lipid metabolism, that is also reflected in *VKORC1* expression levels in murine and human disease.

Several genetic variants in the *VKORC1* locus have been described to influence patients’ response to warfarin treatment. (35,36) However, to the best of our knowledge, no genetic variants in this locus have been described to influence NAFLD. By using a composite variable as a proxy for NAFLD we may capture more of the genetic variability contributing to NAFLD, than when using single surrogate variables. We may capture more of the metabolic phenotype of NAFLD than if only ALT levels had been used, since the NAFLD-S is made up of liver enzymes, biochemical, and anthropometric variables that are highly correlated with NAFLD, obesity and diabetes. Interestingly, GWAS for BMI, TG LDL, total cholesterol and have identified genetic signals in the *VKORC1* locus, and we find strong colocalization signals for *VKORC1* in the liver (***Supplementary Figure 8***). Moreover, human PheWAS data show a strong association with BMI, TG and HDL (among other cardiometabolic values) between variants in *VKORC1*, including a splice donor variant (rs2884737). These data support our transcriptional data that show a dysregulation of lipid metabolism genes upon *VKORC1* knockdown, and our protein-protein interaction network suggests a role for *VKORC1* in lipid and cholesterol metabolism.

Collectively, present, and previous data provide a potential rationale for the involvement of *VKORC1* in the pathogenesis of NAFLD through the regulation of lipid accumulation and cholesterol metabolism in human hepatocytes. To the best of our knowledge, we provide the first experimental evidence suggesting *VKORC1* as a NAFLD susceptibility gene.

In summary, we have expanded our knowledge on the genetic susceptibility for NAFLD by using GWA-and genetic colocalization studies of surrogate markers of NAFLD. Above all, we have established a functional genomic framework to study putative NAFLD genes at scale. Large-scale CRISPRi screens have not only paved the way to study genes involved in various cardiometabolic phenotypes (37), but also intricate multidimensional gene cellular functions. (8) Our efforts have implicated the *VKORC1* gene in the pathogenesis of NAFLD. Taken together, this study provides a sound rationale for use of CRISPRi screens to delineate the roles of known and new putative causal risk genes for both NAFLD, and other cardiometabolic traits.

## Limitations

This work was conceived and primarily executed before the change in nomenclature from NAFLD to MASLD. The authors have after consideration opted to retain the old nomenclature in this manuscript as data referenced herein were generated under these guidelines.

## Supporting information

Main tables and supplementary tables

Supplementary table 8, attached separately.

## Data Availability

All data produced in the present study are available upon reasonable request to the authors.

## Acknowledgements

The authors would like to express their gratitude to all funding bodies contributing to this work, as well as Drs. Frank Chenfei Ning and Annelie Falkevall for generously providing liver cDNA samples from mice on high fat and chow diet.

---

**Supplementary Figure 1.**
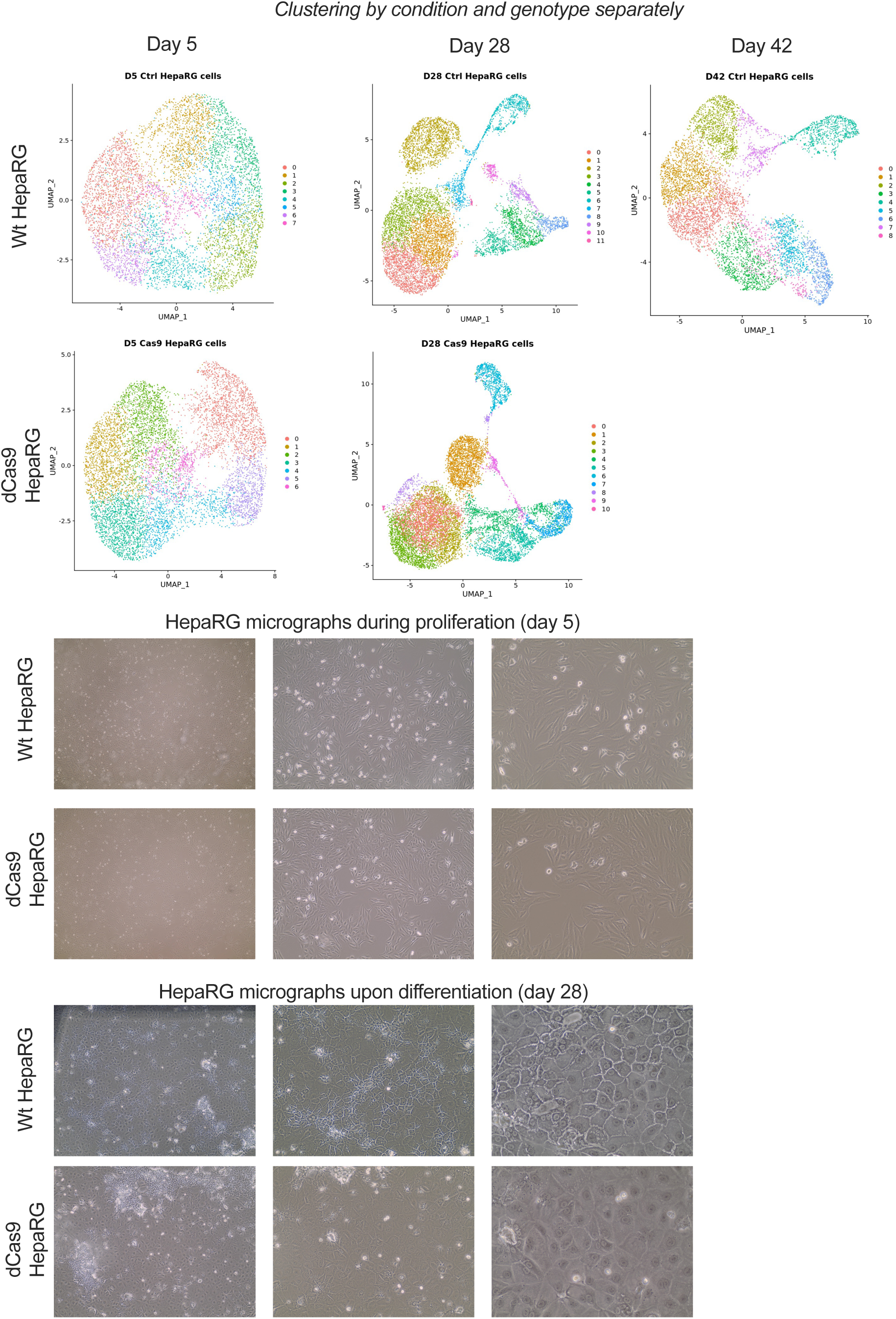
Clustering of HepaRG scRNA-seq characterization experiments. Each genotype and culturing day along the differentiation process was plotted separately. Notably, we observe no major differences in the clustering by genotype or differentiation day (28 days or 42 days post seeding) as shown by Figure 3B. Micrographs showing the gross phenotype of differentiated HepaRG, as has been previously described.

**Supplementary Figure 2.**
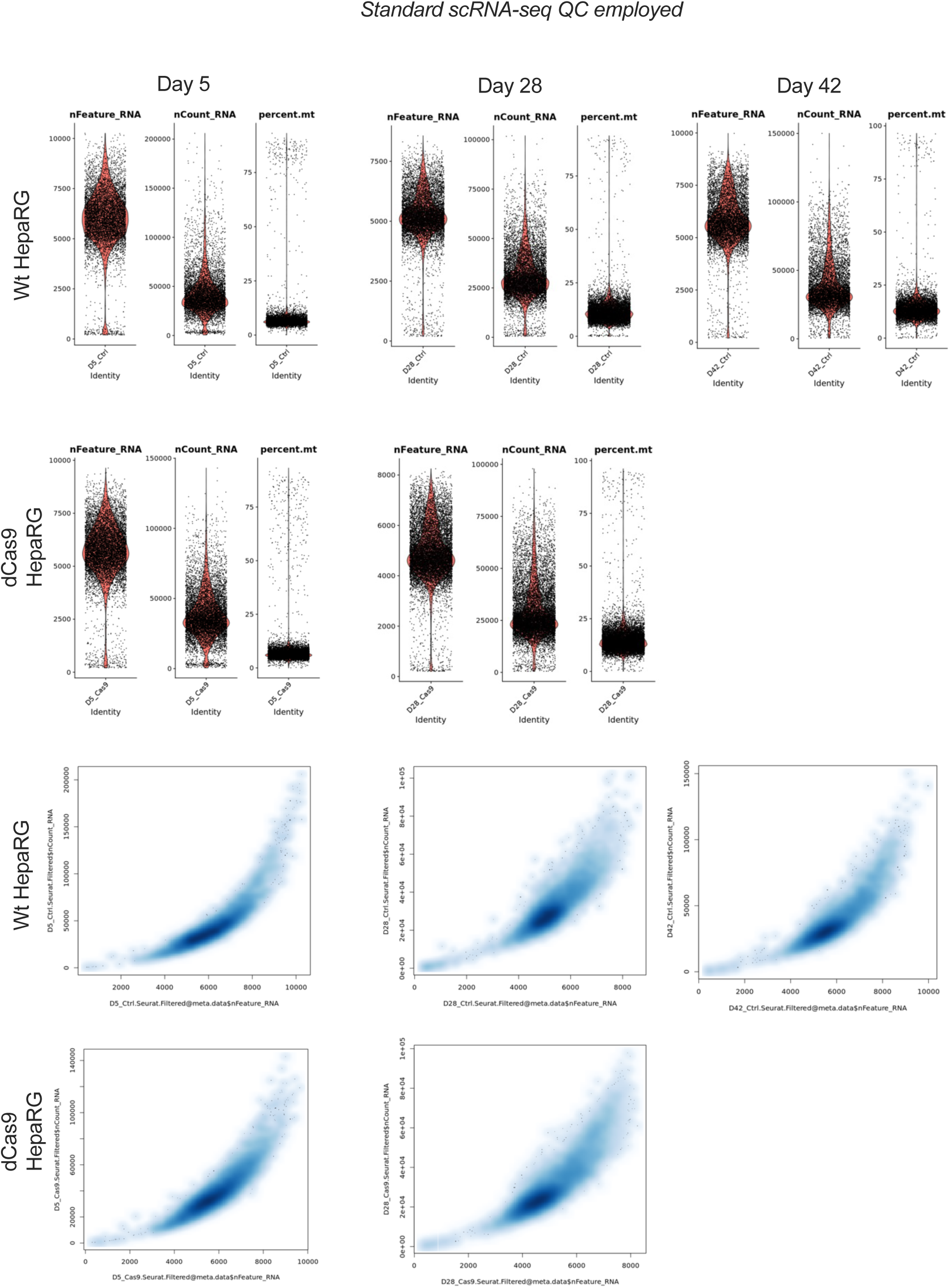
Standard scRNA-seq quality control. Cells exhibiting less than 200 detected features, 2000 RNAs and more than 25% mitochondrial genes (justified based on high expression of mitochondrial genes in differentiated HepaRG cells) were filtered out.

**Supplementary Figure 3.**
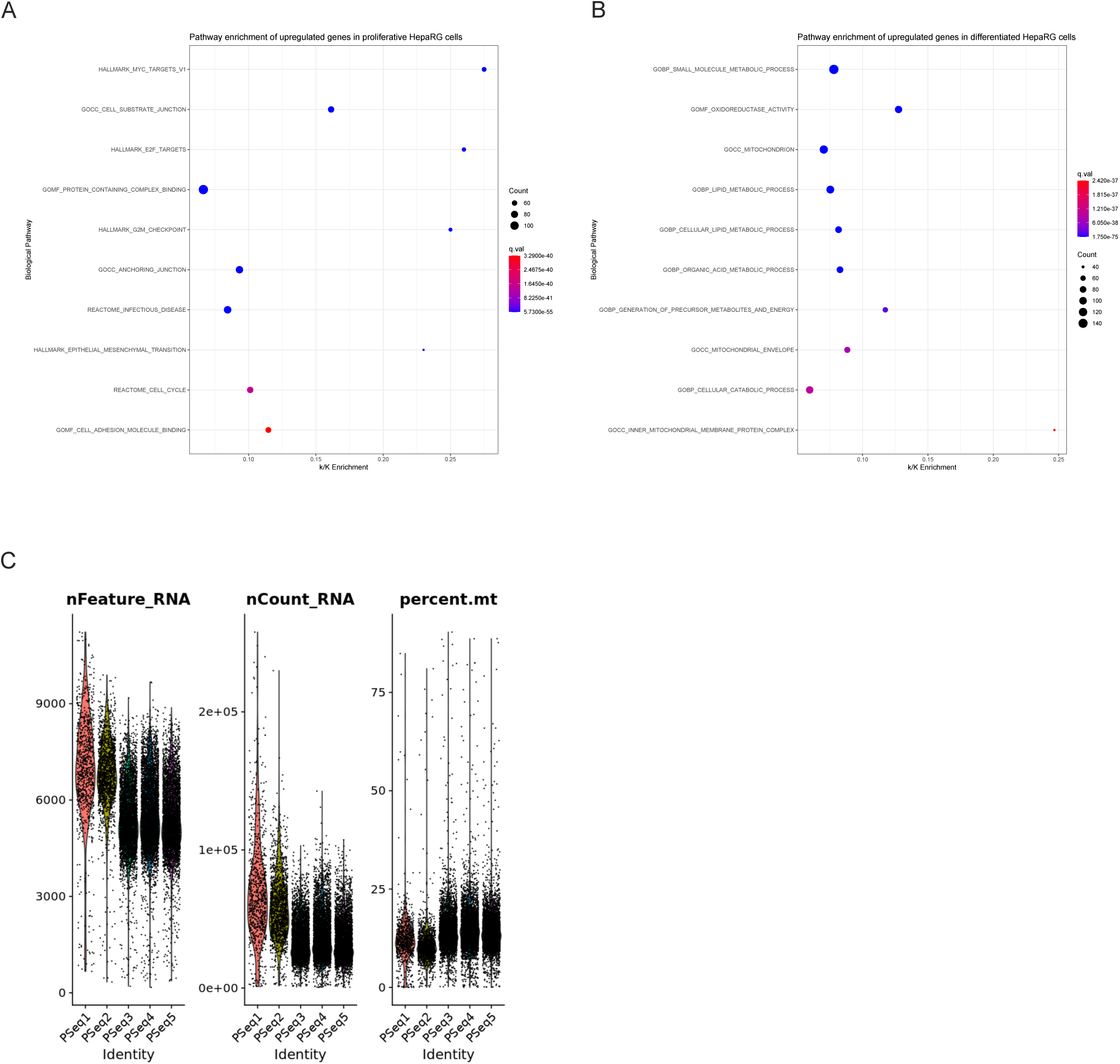
Standard scRNA-seq quality control was carried out on HepaRG cells undergoing Perturb-seq. As before, cells exhibiting less than 200 detected features, 2000 RNAs and more than 25% mitochondrial genes were filtered out.

**Supplementary Figure 4.**
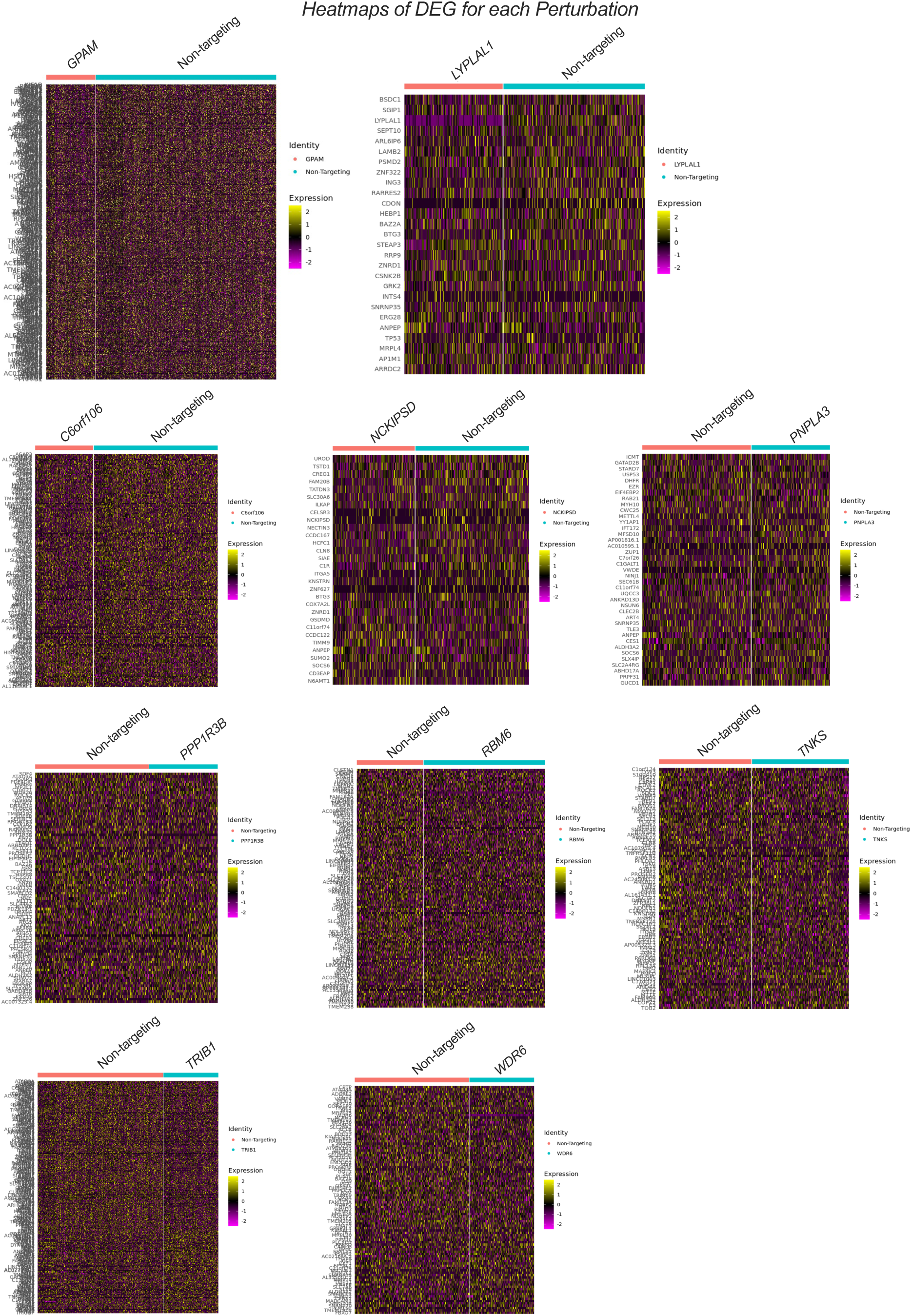
Heatmaps of differentially expressed genes by all targets in Perturb-seq experiments. *GPAM* and *LYPLAL1* are the two top targets after *VKORC1* based on their effects on lipid accumulation. However, gene expression profiles suggest a more striking phenotype of *VKORC1* knockdown compared to all other perturbations.

**Supplementary Figure 5.**
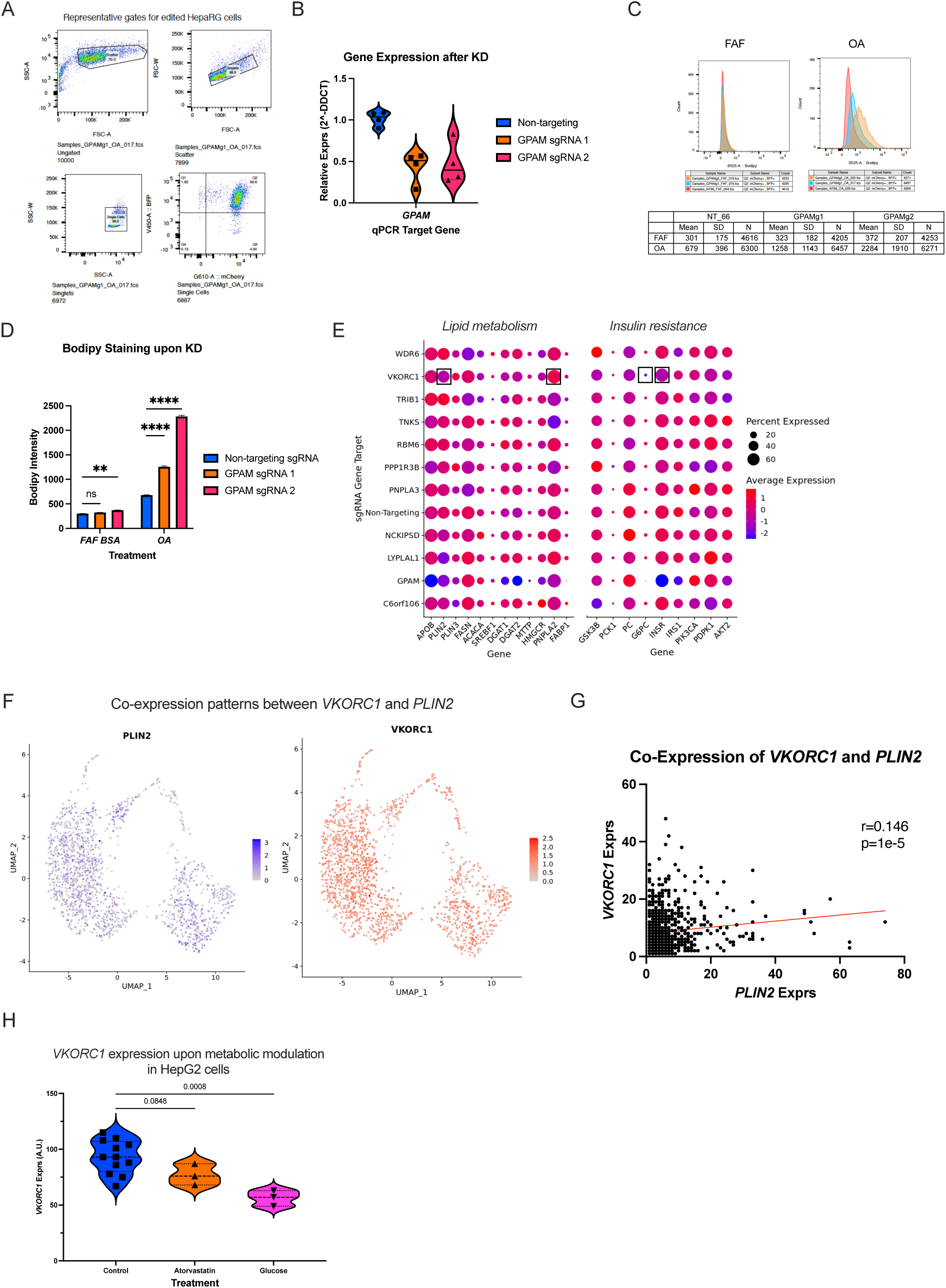
To further validate findings from lipid accumulation-based CRISPRi screens, we preformed single sgRNA transductions targeting the second most interesting target; *GPAM*. **A)** Visualizes FACS sorting gates, where gene-edited mCherry/BFP^+/+^ where sorted. **B)** *GPAM* knockdown was efficient as confirmed by RT-qPCR against the target gene in mCherry/BFP^+/+^ cells, and **C-D)** *GPAM* knockdown increased neutral lipid content in HepaRG cells upon lipid loading. FAF; fatty acis-free BSA, OA; oleic acid. **E)** Dotplot visualizing results of agnostic differential gene expression of *VKORC1* knockdown. Results reveal a trend for downregulation of *PLIN2*, *PNPLA2*, *G6PC*, and *INSR* transcripts in response to *VKORC1* downregulation. **F-G)** Feature plots of single-cell RNA-seq data from cells receiving a non-targeting sgRNA reveal a degree of correlation between the expression of *VKORC1* and *PLIN2*, which reinforces the notion that *VKORC1* may indeed influence the hepatocyte lipid metabolism and thus NAFLD. **H)** Metabolic perturbations in HepG2 cells, revealed that *VKORC1* expression, measured by bulk RNA-seq, is modulated by treating HepG2 cells with glucose, and atorvastatin. Glucose treatment reduces *VKORC1* expression, whereas atorvastatin treatment shows a trend towards lower *VKORC1* expression. *Experimental data consists of n of 3-12 replicates, ANOVA tests were used to explore singificances*. *\* p<0.05, ** p<0.01, ***p<0.001, ****p<0.0001*.

**Supplementary Figure 6.**
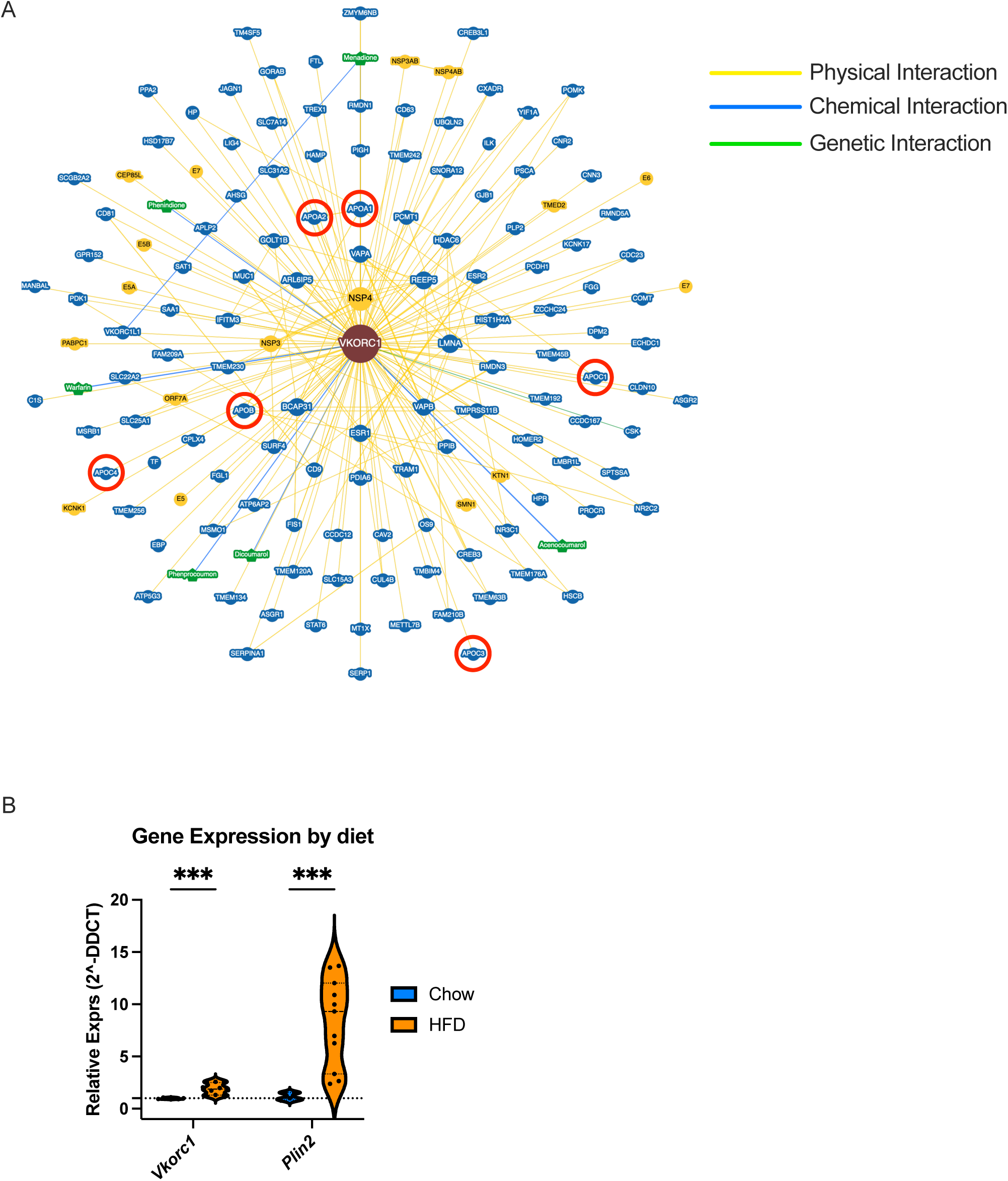
A) Protein-protein interaction network constructed using BioGRID demonstrates a physical interaction between VKORC1 and apolipoproteins, which further reinforces the notion that *VKORC1* may influence lipid and sterol metabolism in liver. **B)** Mice fed a high fat diet, known to induce hepatic steatosis, demonstrate a higher expression of *Vkorc1* along with increased *Plin2* mRNA. *N for experimental data is 6-11 replicates*. *\* p<0.05, ** p<0.01, ***p<0.001, ****p<0.0001*.

**Supplementary Figure 7.**
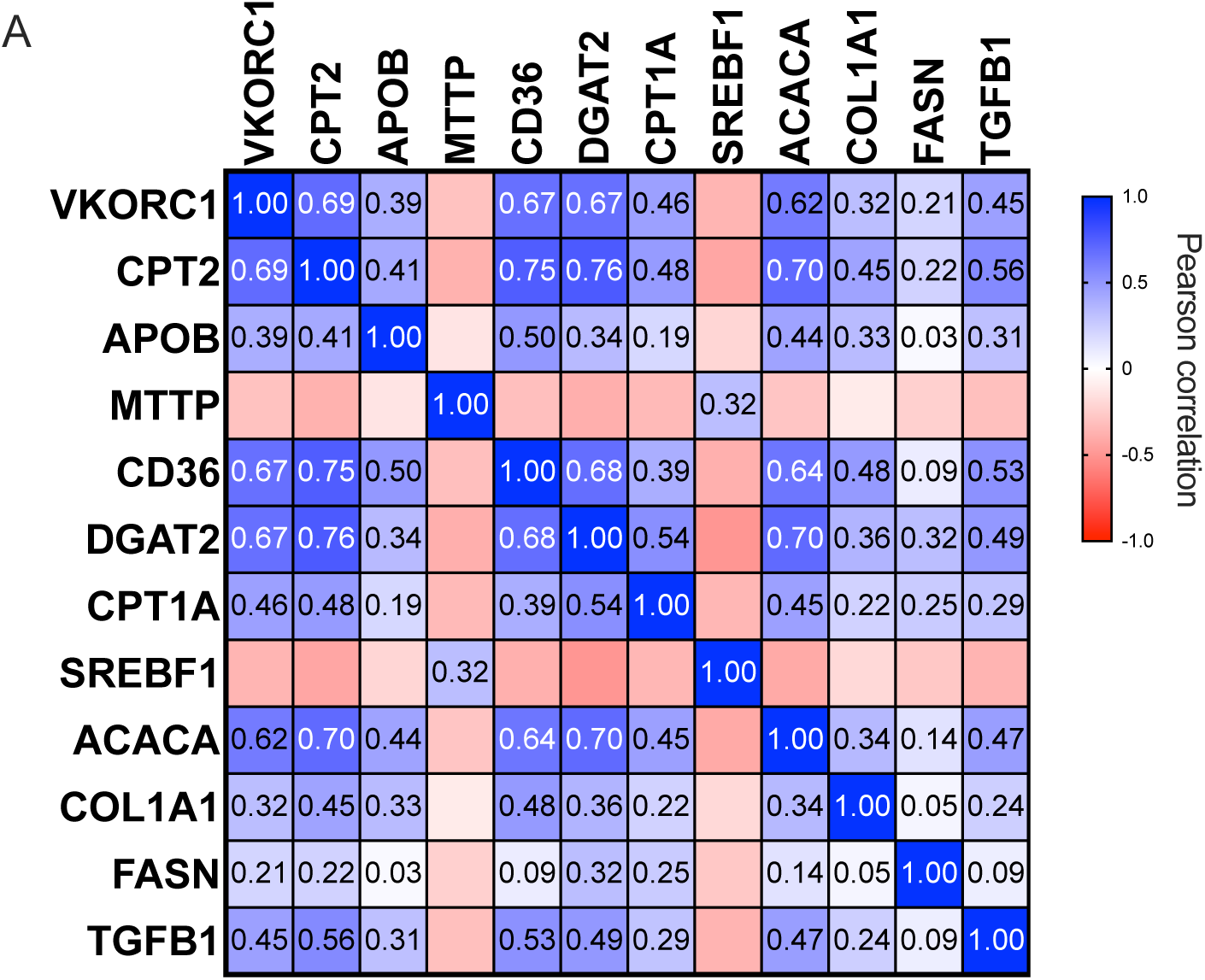
Exploration of *VKORC1* mRNA co-expression, and transcript levels’ relation to biochemical markers of cardiometabolic disease in healthy human liver (the ASAP study). **A)** Co-expression patterns between the *VKORC1* transcript and transcripts involved in the NAFLD pathogenesis. Data reveal that there is a significant co-expression between *VKORC1* and regulators of NAFLD development, which reinforeces the connection between *VKORC1* expression and the early development of NAFLD. *N for co-expression analyses is 210*.

**Supplementary Figure 8.**
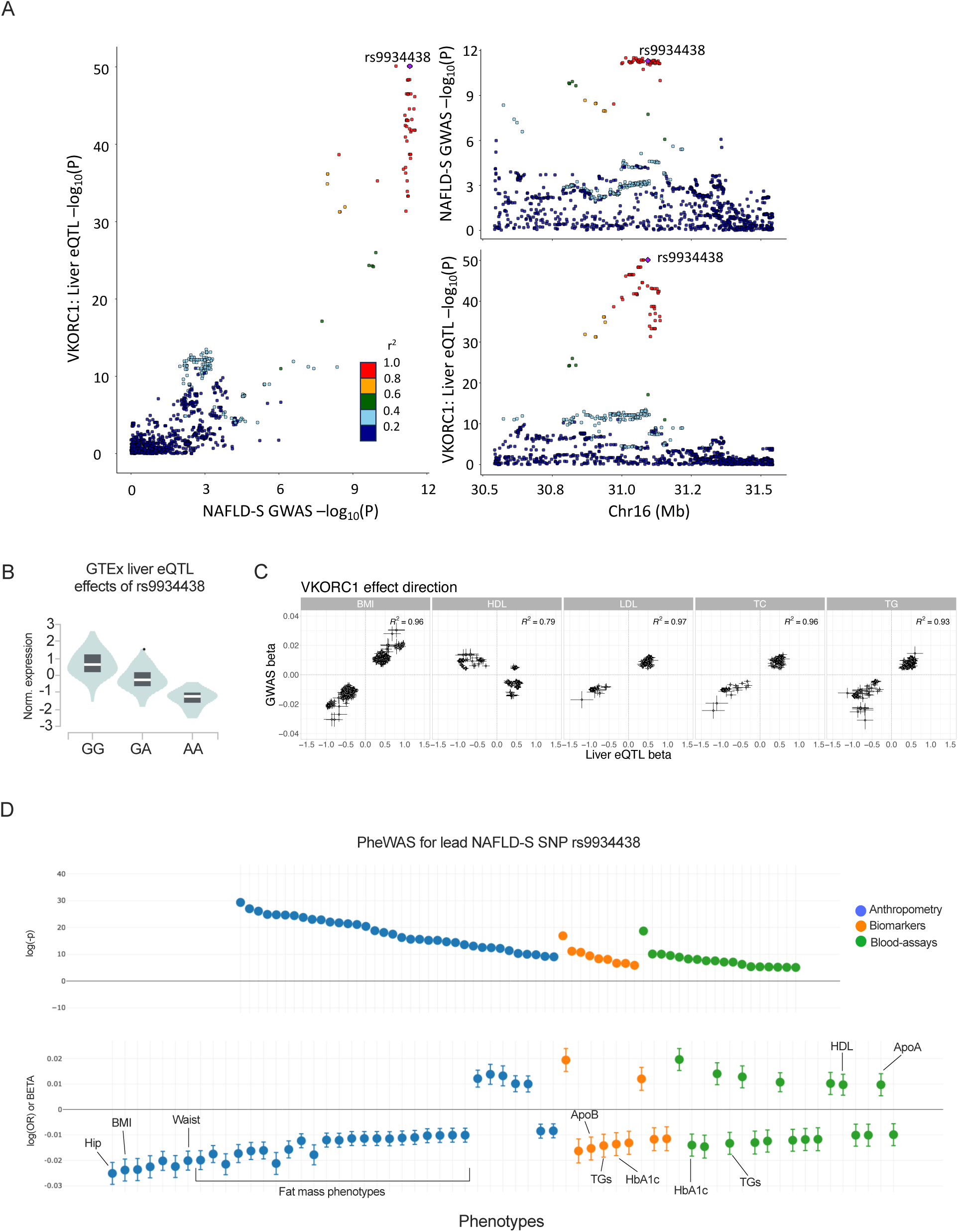
Exploration of the *VKORC1* locus, and associations with other cardiometabolic traits using PheWAS. **A)** Correlation plot between NAFLD-S GWAS p-values and GTEx (v8) liver eQTL p-values, as well as locus plots for NAFLD-S associations and eQTL effects plotted separately, in SNPs used for VKORC1 colocalization. **B)** PheWAS analysis of the NAFLD-S lead SNP rs9934438, and its association with cardiometabolic traits. Notably, the rs9934438 A allele is associated with lower hip and waist circumference, lower BMI and lower values for several fat mass phenotypes in the Biobank engine powered by Stanford University. Furthermore, the rs9934438 A allele is also associated with lower levels of biomarkers of cardiometabolic disease (ApoB, TG, HbA1c), and higher levels of protective biomarkers (HDL, ApoA). **C)** Importantly, the rs9934438 A allele that is associated with protection from NAFLD-S and cardiometabolic biomarkers, and is associated with more protective biomarkers of cardiometabolic disease is also associated with lower *VKORC1* expression. This suggests that lower *VKORC1* expression in the liver may protect from protective of NAFLD and cardiometabolic disease.

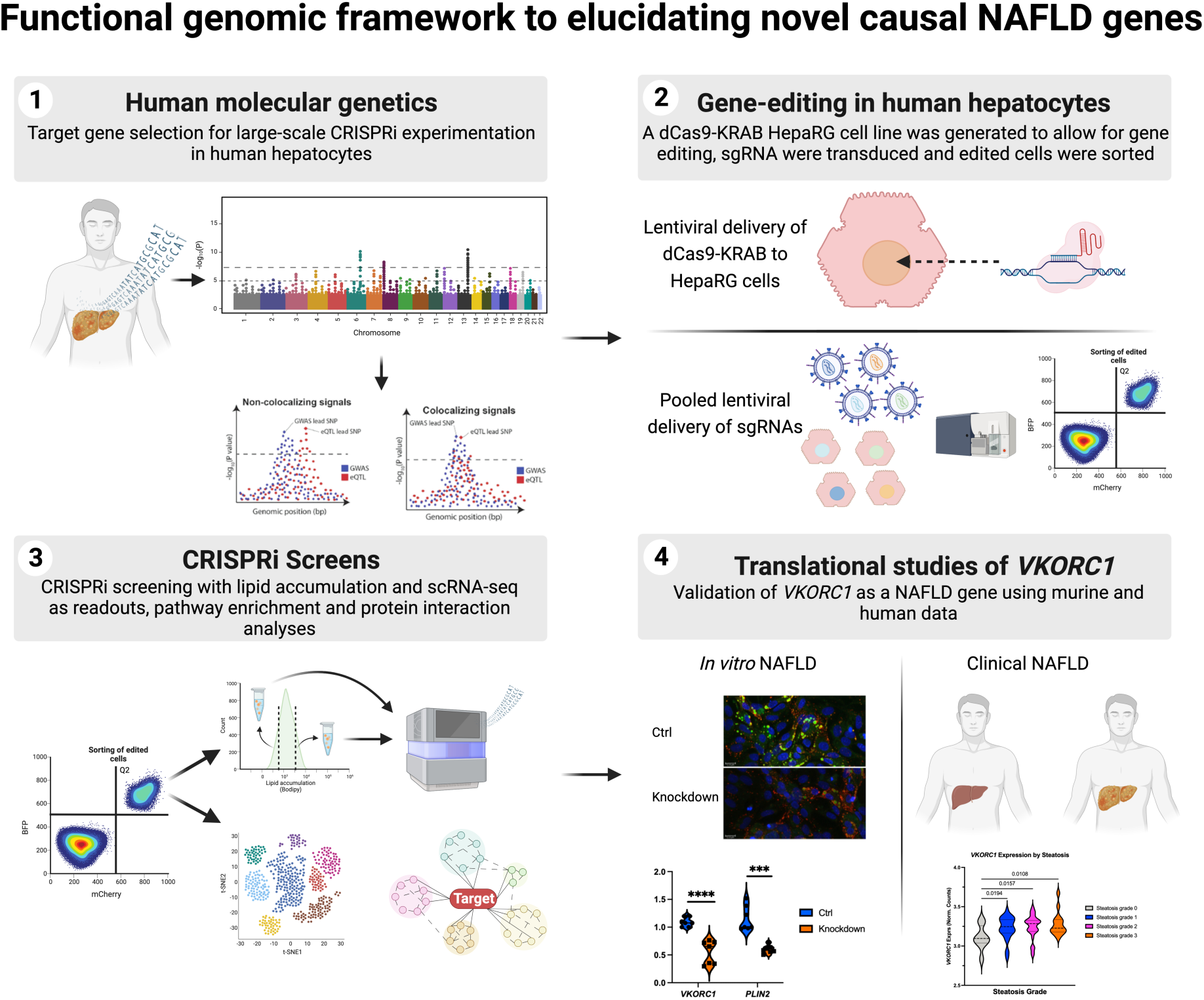

## Notes

**Financial Support:** PSG is supported by the Swedish Research Council (Vetenskapsrådet), grant number 2018-06580 and the Swedish Heart-Lung Foundation grant number 20170221. JMJ is funded by grants from the Novo Nordisk Foundation and the Stanford Bio-X Program (NNF17OC0025806). JWK is supported by the NIH through grants: P30 DK116074 (to the Stanford Diabetes Research Center), R01 DK116750, R01 DK120565, R01 DK106236; and by the American Diabetes Association through grant 1-19-JDF-108. AR is supported by the Finnish Foundation for Cardiovascular Research, Diabetes Research Foundation, Emil Aaltonen Foundation, Ida Montin’s Foundation, Biomedicum Helsinki Foundation, Orion Research Foundation and the Finnish Medical Foundation. The ASAP study was supported by the Swedish Research Council grant number 2020-01442, the Swedish Heart-Lung Foundation grant number 20180451 and a donation by Mr. Fredrik Lundberg. PA is supported by MCIU/AEI/FEDER, UE (PID2021-124425OB-I00) and Basque Government, Department of Education (IT1476-2).

### Competing Interest Statement

The authors have declared no competing interest.

### Funding Statement

PSG is supported by the Swedish Research Council (Vetenskapsradet), grant number 2018-06580 and the Swedish Heart-Lung Foundation grant number 20170221. JMJ is funded by grants from the Novo Nordisk Foundation and the Stanford Bio-X Program (NNF17OC0025806). JWK is supported by the NIH through grants P30 DK116074 (to the Stanford Diabetes Research Center), R01 DK116750, R01 DK120565, R01 DK106236; and by the American Diabetes Association through grant 1-19-JDF-108. AR is supported by the Finnish Foundation for Cardiovascular Research, Diabetes Research Foundation, Emil Aaltonen Foundation, Ida Montins Foundation, Biomedicum Helsinki Foundation, Orion Research Foundation and the Finnish Medical Foundation. The ASAP study was supported by the Swedish Research Council grant number 2020-01442, the Swedish Heart‐Lung Foundation grant number 20180451 and a donation by Mr. Fredrik Lundberg. PA is supported by MCIU/AEI/FEDER, UE (PID2021-124425OB-I00) and Basque Government, Department of Education (IT1476-2).

### Author Declarations

The UKB study was approved by the Northwest Multi-Center Research Ethics Committee and all participants provided written informed consent to participate.

